# Estimating Between Country Migration in Pneumococcal Populations

**DOI:** 10.1101/2023.11.15.23298520

**Authors:** Sophie Belman, Henri Pesonen, Nicholas J. Croucher, Stephen D. Bentley, Jukka Corander

**Affiliations:** Parasites and Microbes, Wellcome Sanger Institute, Hinxton, Cambridgeshire, UK; Oslo Centre for Biostatistics and Epidemiology, Oslo University Hospital, Oslo, Norway; Department of Biostatistics, University of Oslo, Oslo, Norway; Helsinki Institute for Information Technology HIIT, Department of Mathematics and Statistics, University of Helsinki, Helsinki, Finland; MRC Centre for Global Infectious Disease Analysis, Department of Infectious Disease Epidemiology, School of Public Health, White City Campus, Imperial College London, London, W12 0BZ, UK

## Abstract

*Streptococcus pneumoniae* (the pneumococcus) is a globally distributed, human obligate opportunistic bacterial pathogen which, although often carried commensally, is also a significant cause of invasive disease. Apart from multi-drug resistant and virulent clones, the rate and direction of pneumococcal dissemination between different countries remains largely unknown. The ability for the pneumococcus to take a foothold in a country depends on existing population configuration, the extent of vaccine implementation, as well as human mobility since it is a human obligate bacterium. To shed light on its international movement, we used extensive genome data from the Global Pneumococcal Sequencing (GPS) project and estimated migration parameters between multiple countries in Africa. Data on allele frequencies of polymorphisms at housekeeping-like loci for multiple different lineages circulating in the populations of South Africa, Malawi, Kenya, and The Gambia were used to calculate the fixation index (*F_st_*) between countries. We then further used these summaries to fit migration coalescent models with the likelihood-free inference algorithms available in the ELFI software package. Synthetic data were additionally used to validate the inference approach. Our results demonstrate country-pair specific migration patterns and heterogeneity in the extent of migration between different lineages. Our approach demonstrates that coalescent models can be effectively used for inferring migration rates for bacterial species and lineages provided sufficiently granular population genomics surveillance data. Further it can demonstrate the connectivity of respiratory disease agents between countries to inform intervention policy in the longer term.

## 1 Introduction

Inferring migration events in natural bacterial populations between geographically separated regions is generally challenging since bacteria cannot be tagged in the same way as, for example, animals. For bacteria which colonize humans the rate of migration will be a complicated function of the host mobility and ecological factors influencing the success of onward transmission, such as the level of hygiene, use of antibiotics and vaccination campaigns in the host populations. Progress identifying large-scale migration patterns among bacteria has been primarily made for species or strains more likely causing acute infections and serious illnesses, such as cholera Domman et al. [2017], Okoro et al. [2012], Comas et al. [2013], Lassalle et al. [2023]. However, genetic epidemiology has also been used to elucidate the spread of multi-drug resistant (MDR) strains of *Streptococcus pneumoniae* (the pneumococcus) across different global regions. For such commonly asymptomatic bacteria these studies are highly reliant on comprehensive sampling van Tonder et al. [2015], Quintero Moreno and Araque [2018], Croucher et al. [2014]. Overarchingly, variable sampling strategies between countries, poor approximation of between country mobility, and the large time scales of between country pathogen spread can hinder definitive estimates of the weight and direction of between-country spread.

Global genomic sampling as part of the Global Pneumococcal Sequencing (GPS) Project demonstrates that some lineages and serotypes of this bacterium circulate locally, while others are spread globally. With the exception of the aforementioned MDR strains, the time scales of this spread and the frequency, or direction of migration between specific countries leading to the extant lineage distributions remains unclear Gladstone et al. [2019]. While we can use time resolved phylogenies to infer that genome pairs from countries in South Africa and countries elsewhere in Africa become increasingly similar with increasing divergence times, this is still, overall, much less likely than pair similarity within country Belman et al. [2023]. Informing coalescent models with true case count data can reduce the impact of geographic sampling bias, but for an endemic, often asymptomatic pathogen this remains difficult Layan et al. [2023]. Our previous work used human mobility data from Meta Maas [2019] to build a model inferring bacterial movement Belman et al. [2023], but could provide limited information about the direction of spread. Further, reliable between country human mobility data is scarce for continents such as Africa, so similar approaches have limited use in this context Deutschmann et al. [2022], Gabrielli et al. [2019].

In this work we quantify between-country pneumococcal migration among four African countries. Due to the lack of direct observations of between-country migration we are unable to use typical Bayesian techniques. However, population genomics based surveillance data from multiple countries provides an opportunity to consider quantification of migration rates via coalescent models and likelihood-free inference (LFI) Aeschbacher et al. [2013], Wegmann et al. [2009]. In particular we use Approximate Bayesian Computation (ABC), which is a type of LFI in which we compare population summary statistics between simulated and real genomic data to determine (in this case) patterns of pneumococcal migration.

We can explore the impact of different evolutionary parameters on population samples using software packages which simulate coalescing populations Kelleher and Lohse [2020], Kern and Schrider [2016], Ewing and Hermisson [2010]. ms prime is an adaptation to the classical neutral ms simulator which includes demographic parameters such as population size and migration Hudson [2002]. Such a simulation-based inference framework Sisson et al. [2018] enables inference of the unknown migration parameters without access to a closed form expression for the data under any particular set of parameters assumed to govern the migration process. Given the generality of the problem of migration quantification for microbes, our approach is of wider interest beyond the pneumococcal case considered in detail here.

## 2 Materials and methods

### 2.1 Population Divergence Summary Statistics

Rather than comparing the entire nucleotide diversity of genes and genomes within and between populations one can summarize the diversity using relevant statistics derived from population genetics theory under neutrality.

The fixation index (*F_st_*) was first developed by Sewall Wright Wright [1949] in 1949 to describe randomly drawn alleles in one population as compared to the total sampled population. There have been many adaptations to this but the Weir & Cockerham (WC) *F_st_* Weir and Cockerham [1984] is the most commonly used today and describes the total population as the most recent ancestral population. The WC *F_st_* is a parameter describing the evolutionary process of drift rather than a statistic of observed samples and assumes equal drift across populations. When the *F_st_* for each population is different the WC estimator becomes a function of the ratio of sample sizes between them rather than true divergence. Further WC *F_st_* suffers from the ‘star phylogeny’ assumption in that all populations independently descended from the same ancestor. Weir & Hill adapted the WC *F_st_* to estimate population specific values and Hudson et al. adapted this to estimate the *F_st_* between populations in 1992 Hudson et al. [1992], Selander and Hudson [1976], Holsinger and Weir [2009]. Hudsons *F_st_* is more robust to variable sample sizes and variable *F_st_* values across populations Bhatia et al. [2013]. Simply Hudsons *F_st_* can be described by Weir & Hills single population *F_st_* estimate.

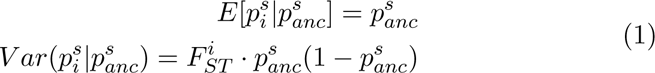

where 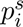 is the allele frequency in population *i* at SNP *s*, and 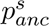 is the frequency of the same allele at that SNP *s* in the ancestral population and 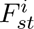 is the population-specific *F_st_* for population *i*. Hudsons estimate combines these to estimate the *F_st_* for a pair of populations to be

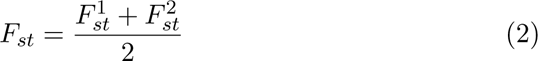

where 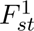 is population 1 and 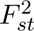 is population 2.

### 2.2 Inference Strategy Overview

#### 2.2.1 Simulator

We used msprime Kelleher and Lohse [2020], Nelson et al. [2020], Baumdicker et al. [2022] as the simulator in this framework. msprime is a software package which simulates the coalescent process for thousands of genomes. It can incorporate recombination, mutation, and migration between demes. It outputs phylogenetic trees representing the population, onto which mutations can be imputed and summary statistics calculated. These summary statistics are then compared to summary statistics from the real data. The coalescent simulation requires specific parameters. For the two deme model we included geographic demes *A* and *B* and initial population size for each deme (*P_A_*, *P_B_*). The sample sizes drawn from each population (*samp_A_*, *samp_B_*) are included as parameters and crucially, we include the inferrable parameters for the migration rates asymmetrically from deme *A* to deme *B* (*mig_A,B_*), and the inverse (*mig_B,A_*). We also include a mutation rate (*θ*). We specified these parameters scaling the mutation rate to the length of genome we input and down-sampled the true population sizes from each country for computational efficiency. We included the infinite sites model in this framework which rather than allowing only a finite number of mutations per-site allows an infinite number across continuous space whereby no site mutates twice Kimura [1969], Ma et al. [2008] (Table 1). We employ replicates, both for the simulator and the real-data, to reduce variability around the summary statistic estimate (Table 1).

**Table 1:** Parameter summary for inference validation. Estimates of true parameter values for the deme population size, mutation rate in sites/per-year, whether there is a discrete genome, and sequence length for left) truth and right) within inference validation msprime simulation frame-work.

| Parameters | Truth |  | Simulated Input |  |
| --- | --- | --- | --- | --- |
| Deme Population Sizes | South Africa | 60.04 M | South Africa | 6000 |
|  | Malawi | 19.65 M | Malawi | 2000 |
|  | Kenya | 54.99 M | Kenya | 5000 |
|  | The Gambia | 2.487 M | The Gambia | 1000 |
| Mutation Rate ($\theta$) | $1.57E - 06$ | | $2.50E - 05$ | |
| Discrete Genome | True (finite sites model) |  | False (infinite sites model) |  |
| Sequence Length | 2 Mbp |  | 500 |  |
| Migration Rate | ? | | $mig_{ab} = 0.6$ | |
| | | | $mig_{ba} = 0.1$ | |

#### 2.2.2 Parameter Inference Algorithm

LFI methods such as ABC were originally developed to do statistical inference with a very large number of synthetic observations from the simulator, rejecting non-conforming observations and keeping only observations close the observed data Sisson et al. [2018]. However, with a complex process involving multiple unknown parameters millions of simulations are often necessary to infer the required parameter using the basic rejection algorithm. A state-of-the-art ABC method such as the Bayesian Optimization for Likelihood-Inference (BOLFI) algorithm employs active learning to reduce the required computer simulation multiple fold by focusing only on the relevant parts of the parameter space. This can be done using a probabilistic model such as a Gaussian Process to model the relationship of parameters and the discrepancy between the observed data and synthetic data, and seeking to minimize this Gutmann et al. [2016]. BOLFI provides us with a surrogate likelihood function that we use in a Bayesian framework along with the parameter prior distribution to obtain the posterior distribution. In practice, a sample is drawn from the posterior using e.g. MCMC sampling to calculate posterior means and other summarizing estimates.

#### 2.2.3 Model Validation: Recapturing Simulated Data

##### Two Deme Model

To validate this inference framework we first conducted prior predictive analysis to determine the sensitivity of the summary statistic, *F_st_*, to a range of migration parameters. We maintained initial populations sizes for A and B of 6000 and 2000 with 600 sampled from each. Other parameters include a sequence length of 500, *θ* set to 2*e^−^*^5^, and 500 replicates. We divided the migration parameter by 5000 for scaling (Figure 1A). We also evaluated the sensitivity of the *F_st_* to other parameters in the simulator framework including the initial and sampled population sizes, and the mutation rate (Figure S1A-C). We input specific asymmetric migration parameters (*mig_A,B_* and *mig_B,A_*) between two demes (*A* and *B*) and attempted to recapture them using simulated data. For our validation we again set the mutation rate to 2.5 · 10*^−^*^5^, set the number of replicates to 500, sequence length of 500, *samp_A_*=600, *samp_B_*=600, *P_A_*=6000, *P_B_*=2000, corresponding to the population sizes of each country as estimated from LandScan Rose et al. [2020](Table 1). We use the infinite sites model by setting discrete genome to FALSE thus including infinite possible SNP sites. We calculate the *F_st_* for each simulation and use 9 quantiles at 0.1 increments across replicates for the summary statistic. We use uniform priors *mig_A,B_* ∼ Unif[*L, U*] and *mig_B,A_* ∼ Unif[*L, U*] to calculate the posterior distributions where *L* represents the lower bounds and *U* represents the upper bounds. We input 0.6 and 0.1 for *mig_A,B_* and *mig_B,A_* respectively(Figure 1B-C; red line, Figure S2) The estimated parameters for *mig_A,B_* and *mig_B,A_* respectively were 0.207 [95% Confidence Intervals (CIs) 0.011-0.422] and 0.477 (95% CIs 0.164-0.721) (Figure 1B-C; blue dashed line, Figure S2).

**Figure 1:**
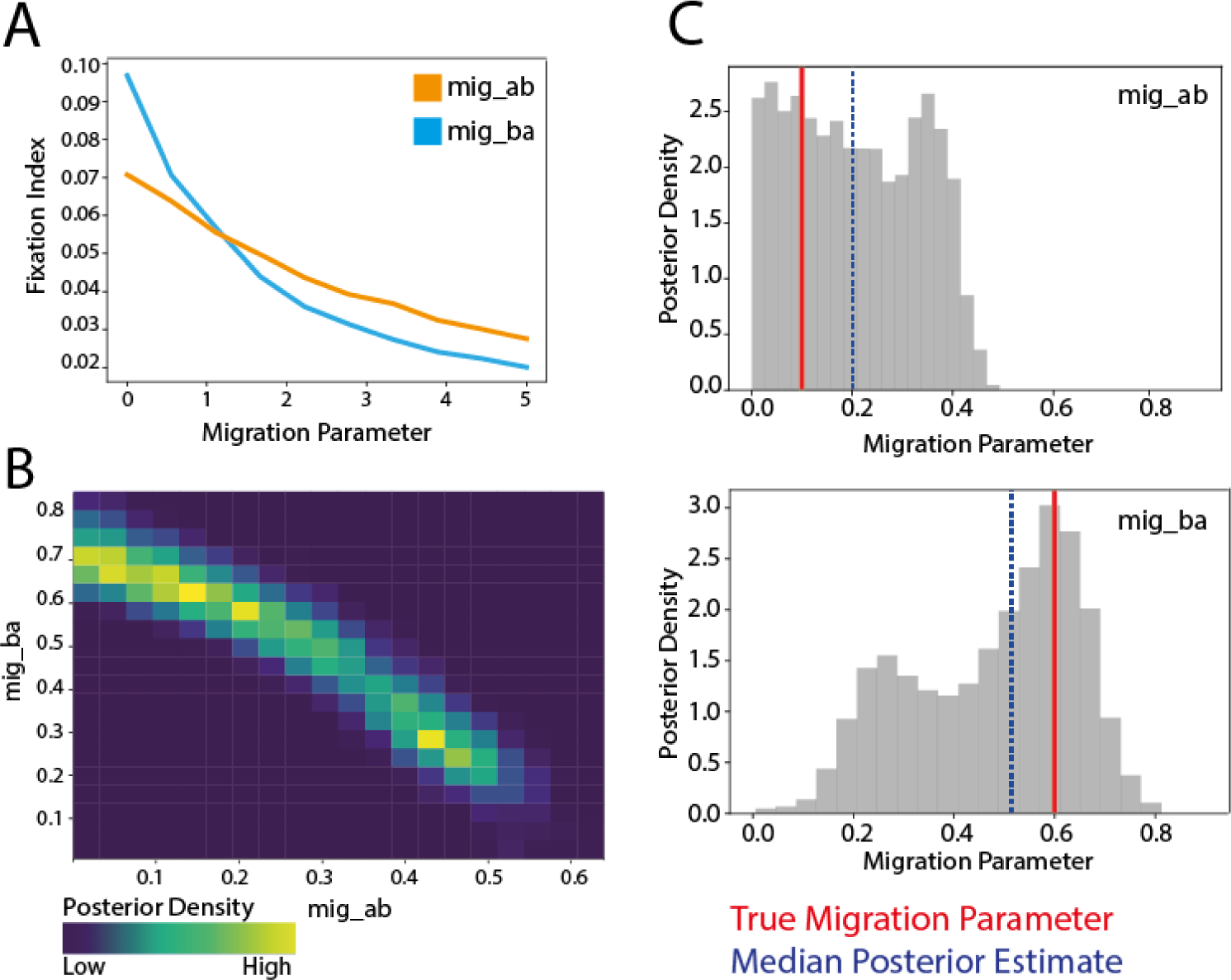
Recapturing input migration parameters with a 2 Deme model. A) Prior predictive analysis testing the sensitivity of our summary statistic, the fixation index, to our parameter estimates for each parameter in an asymmetric model. Initial populations sizes for A and B of 6000 and 2000 with 600 sampled from each. Other parameters include a sequence length of 500, *θ* set to 2e-5, and 500 replicates. The migration parameter is divided by 5000 for scaling. B) The overlapping posterior density migration parameter estimates for migration parameter ‘*mig_ab_*’ — from a population A (initial population size 6000) to population B (initial population size 2000) and ‘*mig_ba_*’ from population B to population A. The ‘true’ input parameters were *mig_ab_*=0.1 and *mig_ba_*=0.6. C) The posterior densities visualized individually for each parameter (grey), the true input parameter is indicated by the red vertical line while the median posterior estimate is indicated by the blue dashed line.

We repeated this validation varying the initial population size of each deme relative the true population size of each country(Table 1, Figure S2). There is co-linearity between the asymmetric parameters. The model is able to resolve this and recapture the correct peak for each parameter.

##### Four Deme Model

We validate a symmetric four deme model in which the weight, not the direction of migration, is estimated between all four deme pairs within one model. We conducted prior predictive analysis to determine appropriate bounds for the uniform prior (Figure S3) and the impact of each parameter on all other parameters. We found that the parameters were not independent of each other and altering one migration parameter impacted the *F_st_* of another (Figure S3) implying that migration between two countries may impact the migration estimates in a third or fourth additional country. This relationship is expected due to the migration across all demes included in the simulation which is inherent to our framework.

To validate the four deme model we input 6 migration parameters (*mig_a_b* = 2.5*, mig_a_c* = 2.5*, mig_a_d* = 2.5*, mig_b_c* = 1.5*, mig_b_d* = 1.5*, mig_c_d* = 0.5) and were able to recapture them within the symmetric model. The true migration parameter is indicated by the red vertical line while the estimated median parameter is indicated by the blue-dashed vertical line. The input population sizes for each of the demes scale to the true population size and are indicated in Figure S4. All other parameters are consistent with those described in Section ‘Two Deme Model’ however only a single migration parameter is input for each deme pair.

### 2.3 Application to a Pneumococcal Dataset

#### 2.3.1 Isolate Culture and Sequencing

We included pneumococcal genomes from four Sub-Saharan African countries including South Africa (N=6919), The Gambia (N=3090), Malawi (N=1612), and Kenya (N=961) from the GPS Project in our initial dataset GPS et al. [2022](Figure 2A, Table 5). The isolates were collected between 1990 and 2014 (Figure 2B), comprised 360 GPSCs and 83 different serotypes, and were randomly selected for sequencing (Figure 2C). *‘Country’* will be used interchangeably with *‘Deme’* throughout this manuscript. We will also interchangeably use *‘GPSC’* and *‘lineage’*.

**Figure 2:**
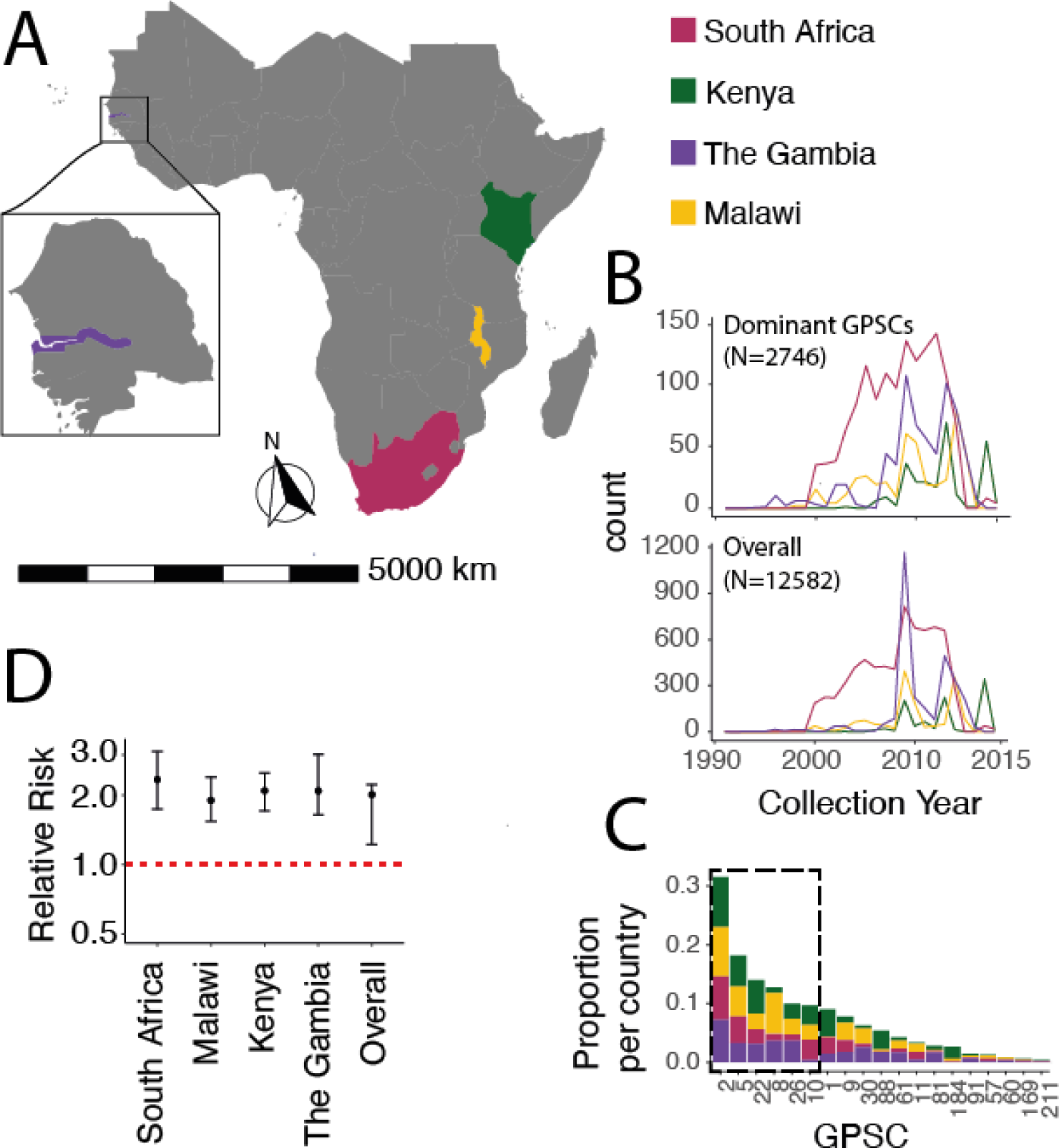
Population composition of four demes. A) Map of isolation location of pneumococcal genomes including South Africa (pink), Kenya (green), The Gambia (purple), and Malawi (yellow). B) Bottom) The isolates were collected from 1990-2014 and the total dataset included N=12582 isolates Top) subsetting by the ‘Dominant GPSCs’ those spanning all four demes and comprising *>*1% of the GPSCs in each country included six total GPSCs highlighted in C) which shows the proportion of total GPSCs each GPSC comprised in each country. The dominant six are outlined in a grey box. D) The relative risk of similarity within country as compared to between countries at the lineage level for each country and the overall risk of being the same GPSC from the same country compared to different countries.

We calculated the relative risk of GPSC similarity by country as per the method described in Belman *et al*. [2023] Belman et al. [2023]. To determine whether there are distinct GPSCs circulating in each country we calculated the risk that a pair of isolates sampled from the same country would be the same GPSC as compared to a pair of isolates selected from each country and another country (Figure 2D). There was a 2.02 (95%CIs 1.23-2.23) fold increased risk of a pair being the same GPSC when from the same country as compared to pairs from different countries indicating that there were distinct GPSCs circulating in each.

These pneumococcal isolates were selectively cultured on BD Trypticase Soy Agar II with 5% sheep blood (Beckton Dickinson, Heidelberg, Germany) and incubated overnight at 37°C in 5% *CO*_2_. Genomic DNA was then extracted manually using a modified QIAamp DNA Mini Kit (QIAGEN, Inc., Valencia, CA) protocol. As part of GPS, pneumococcal isolates were whole-genome sequenced on the Illumina HiSeq platform to produce paired-end reads with an average of 100-125 bases in length and data were deposited in the European Nucleotide Database. Whole genome sequence data was processed as previously described Gladstone et al. [2019].

To control for population structure, and be sure the parameters we were estimating were not just due to lineage diversity in each country, we included only GPSCs which are at greater than 2% prevalence overall in the population (11 GPSCs), we then limited to only those with isolates present in each of the four countries (7 GPSCs), and finally we restricted based on those GPSCs which comprised *>*1% of the remaining number of isolates in each country. Ultimately we included 6 ‘Dominant GPSCs’ (GPSC2, GPSC5, GPSC8, GPSC10, GPSC22, and GPSC26) which included a total of 2746 genomes.

#### 2.3.2 Neutral Gene Selection

We selected genes which are less impacted by evolutionary selection processes due to their less than 0.1 median IgG binding affinity, and ‘non-antibody binding’ status in Croucher *et al*. [2017] Croucher et al. [2017]. The selected genes are present with at least a 99% frequency across our dataset. Across the whole dataset we included two groups of ‘neutral’ genes: 1) a subset of 341 genes which were non-ABT, and 2) 84 genes which had *<*0.1 median IgG binding affinity *and* were non-ABT. For the by-GPSC analysis we selected 81 and 355 ‘neutral’ genes which were core across all six Dominant GPSCs and fit the same criteria as above.

#### 2.3.3 Pairwise Distances Between Genes

We built genome alignments for the previously described sets of genes utilizing a combination of Panaroo Tonkin-Hill et al. [2020] and BioPython Chapman and Chang [2000]. We calculated pairwise distances from every genomes to every other genomes using both Hamming (Figure S5A) Hamming [1950] and Jaccard distances (Figure S5B) Murphy [1996], Jaccard 1912 grouping the genomes by isolation country. If there were distinct, qualitative differences between countries one would expect clear divergent blocks of similarity (lighter color) along the diagonal of the pairwise distance plots. We used scikit-allel Miles et al. [2021] for all pairwise distance calculations.

At the population level (N=12582) the homogenous color across both Jaccard, and Hamming distance plots is representative of mixed populations across the countries. Despite the 2-times higher probability of a pair being the same GPSC when from the same country as compared to pairs from different countries there are still many similar GPSCs between them (Figure S5).

#### 2.3.4 Controlling Population Structure

##### GPSC Level

To control for population structure we interrogated each Dominant GPSC (N=2746), for the pairwise distances between genes across countries. We only included genes which were present in all four countries and across the Dominant GPSCs (N=81: non-ABT & *<*0.1 IgG Binding; N=355: non-ABT). The 81 selected genes had a median gene length of 555 bp (95% CIs 207-1809)(Figure S6).

We grouped the alignments by country to see if there were qualitatively distinguishable differences within versus between country. Again we calculated Hamming distances for 81 and 355 (Figure S7) genes. We repeated this using Jaccard distances for both gene sets (Figure S8). Jaccard and Hamming distances resulted in similar patterns.

##### Linked SNP Sites

Co-selected sites may exacerbate the signal within countries due to recombination, and thus mask the migration signal between countries. To exclude co-selected sites we calculated the mutual information score (MI) across the concatenated neutral genes both overall and for each GPSC. We visualized these using SpydrPick Pensar et al. [2019] to understand the relationship across all 81 genes bi-allelic SNP sites (Table 2, Figure S9). The majority of sites with high, direct MI scores were within 1kb of each other. GPSC2 and GPSC8 have high linkage across many nucleotide distances which span selected genes. These are invasive lineages and undergo less recombination than other lineages. This is demonstrated by each only comprising a single PCV13-type serotype, serotypes 1 and 5 respectively. You can explore each lineage phylogeny at the Microreact web server (Table 4).

**Table 2:** Biallelic SNP count for each GPSC. Including total number of biallelic SNPs, total excluding all within a 1kb window upstream with an *r*^2^ *>* 0.5, and total excluding all within a 1kb window upstream with an *r*^2^ *>* 0.05.

| GPSC | Total SNPs | Excluding 0.5 Threshold | Excluding 0.05 Threshold |
| --- | --- | --- | --- |
| 2 | 968 | 214 | 171 |
| 5 | 1745 | 336 | 161 |
| 8 | 580 | 78 | 62 |
| 10 | 1221 | 176 | 104 |
| 22 | 1469 | 294 | 132 |
| 26 | 543 | 92 | 60 |

Given the distribution of gene length when the distance between SNP sites exceeds approximately 1.8kb the sites are separated by more than one gene length (Figure S6) resulting in ultimately fewer genes being included. We controlled for co-selection between SNPs with a strict correlation threshold minimum of 0.05 *r*^2^ and a more flexible minimum correlation of 0.5 *r*^2^. We used bcftools +prune to remove all SNPs with an *r*^2^ greater than the threshold within a 1kb upstream window and repeated our two deme migration analysis for each GPSC between South Africa and Malawi (Table 2). A 1kb window encompasses the entire length of the majority of genes included (Figure S6). We proceeded with the 0.5 relatedness threshold as it excluded fewer SNP sites but maintained similar estimates as the stricter threshold(Figure S10).

#### 2.3.5 Fixation Index

To quantify the divergence between each location we then calculated *F_st_* overall and by GPSC using tskit Baumdicker et al. [2022], Kelleher and Lohse 2020, Nelson et al. [2020]. We compared the SNP *F_st_* between Weir-Cockerham and Hudsons *F_st_* using tskit within the coalescent simulation software msprime Kelleher et al. [2016], Nelson et al. [2020], Baumdicker et al. [2022]. The Hudson’s *F_st_* is calculated in tskit:

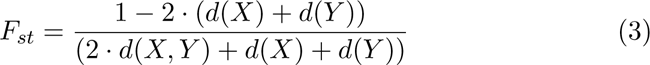

where *X* and *Y* and *d*(*X*) and *d*(*Y*) are the populations and diversity of those populations respectively and *d*(*X, Y*) is the shared diversity of both populations. A higher *F_st_* corresponds to a more divergent, separate population, while a lower *F_st_* corresponds to a more highly mixing population, also known as panmictic.

We compare the *F_st_* estimates from Weir & Cockerham and Hudson and find them to be largely linear with some over-estimates by WC. Due to the uneven sample sizes of our populations we proceed with the Hudson’s estimate (Figure S11).

We calculate the *F_st_* across all genomes between each of the four demes for each GPSC. We repeat this including 81 neutral genes, 355 neutral genes, and the *pbp* genes as a control for selection (Figure S12). We find variable estimates both across GPSCs and between countries. Notably GPSC2 and GPSC8 have higher estimates overall than the rest of the GPSCs in both the 81 and 355 gene comparisons (Figure S12A-B). We included the *pbp* genes as these genes confer resistance to *β*-lactams and are under significant selective pressure. Given variable selective pressures across countries they would be expected to have different cross-country diversity patterns than our ‘neutral’ selected genes. We do see differences between the *pbp* gene divergence as compared to the divergence patterns we see when including the ‘neutral’ genes. This is reassuring as we expect a different pattern as a result of AMR selection. The *F_st_* for the 355 genes was less informative with regards to the different populations. Hereafter we only include the 81 gene analysis by GPSC. All analyses and visualizations was conducted in Python v3.9.13 and R v3.6.1.

### 2.4 Overall Between-Country Migration Risk

While the above framework identifies migration parameters when migration occurs, it does not account for the actual probability of movement between countries as compared to movement within a country. To address this we applied a simple relative risk framework incorporating divergence time between genome pairs. We included those most prevalent GPSCs across the dataset GPSC2 (N=904), GPSC5 (N=473), GPSC10 (N=306). We created reference genomes for each GPSC using ABACAS to order the contiguous sequences (contigs) from a representative of each GPSC mapped to *Streptococcus pneumoniae* (strain ATCC 700669/Spain 23F-1) [EMBL accession: FM211187]. Any contigs which did not align were concatenated to the end. We multiply mapped all genomes from each GPSC against these references respectively using a custom mapping, variant calling, and local realignment around indels pipeline using bwa-MEM Li and Durbin [2009] and samtools mpileup Li et al. [2009]. We built trees masking recombination regions using Gubbins Croucher et al. [2015] with RAxML Stamatakis [2014] and a general time reversible (GTR) evolutionary model. We converted branch length to time using BactDating with a mixed gamma, relaxed clock model Didelot et al. [2018].

We compare the location (*loc*) and label (*G*) (genetic similarity) of pairs of sequences (*i*,*j*) that were collected around the same time (*t*). This approach has been shown to be robust to substantial biases in timing and location of isolate collection. To determine at what divergence time it became equally likely that a pair of genomes were within the same country as between different countries, for each country, we constructed pairwise matrices comparing every isolate to every other isolate (N Pairs=1683). We then determined the proportion of genomes at each divergence time across rolling 10-year time windows within a country as between countries. Dividing the proportion which are within the same country by the proportion between countries gives the relative risk.

In this case the numerator contains the ratio of pairs which are at each divergence time, collected within 10 years of each other *t*, from the same country *loc*, over the total number of pairs collected within 10 years of each other *t* ≤ 10*years* from the same country. The denominator is the ratio of pairs which are within each divergence time *G*, collected within 10 years of each other, from different countries (*L_ref_*) over the total number of pairs collected within 10 years of each other from different countries. Geographic distances were calculated based on the centroid coordinates of each province (Equation 4).

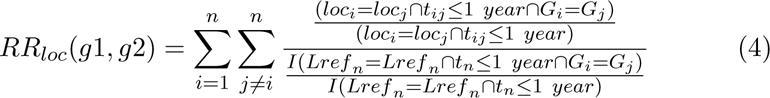

To quantify uncertainty, we used a bootstrapping approach where in each bootstrap iteration we randomly sampled with replacement the isolates before recalculating the statistic. We report the 2.5 and 97.5 percentiles from the resulting distribution.

## 3 Results

### 3.1 Inference Strategy

In brief, to estimate between country migration parameters we developed a framework which uses summary statistics to characterize a sampled pathogen population and compares them to corresponding statistics from a simulated pathogen population under a given coalescent model. We use the Hudsons fixation index (*F_st_*) as the summary statistic in this model. We employ several key pieces of software including msprime to simulate a coalescing population, and the Engine for Likelihood-free Inference (ELFI) Lintusaari et al. [2018] to compare the simulated populations with the observed population data. Broadly, our strategy falls under the Approximate Bayesian Computation (ABC) paradigm Sisson et al. [2018].

Using this framework we develop two models for quantifying migration. The two-deme model compares pairs of countries and determines the asymmetric migration parameters between them (the amount of migration from country A to country B and the reverse). The other model is a four-deme model quantifying symmetric migration parameters between four countries, encapsulated in six rate parameters.

### 3.2 Application to a Pneumococcal Dataset

We implement both of these models using genomes from the GPS project (Figure 2A, Table 5) GPS et al. 2022. We included GPSCs (also referred to as lineages throughout this paper) representing pneumococcal between-country variation in neutral genes and approximately un-linked SNP sites. We included 12582 genomes for initial exploration but ultimately reduced this to 6 ‘Dominant GPSCs’(GPSC2, GPSC5, GPSC8, GPSC10, GPSC22, and GPSC26) (N=2746 genomes) from South Africa, The Gambia, Malawi and Kenya for the migration models.

The isolates were collected between 1990 and 2014 (Figure 2B), comprised 360 GPSCs and 83 different serotypes, and were randomly selected for sequencing (Figure 2C). *‘Country’* will be used interchangeably with *‘Deme’* throughout this manuscript. We will also interchangeably use *‘GPSC’* and *‘lineage’* (Figure 2B-C, Table 3, Table 4).

**Table 3:** The number of genomes for each country and GPSC. Including only GPSCs which were *>*2% prevalence overall, present in all four countries, and present at >1% prevalence in each country.

|  | GPSC10 | GPSC2 | GPSC22 | GPSC26 | GPSC5 | GPSC8 | Total |
| --- | --- | --- | --- | --- | --- | --- | --- |
| South Africa | 232 | 506 | 173 | 73 | 309 | 82 | 1376 |
| Malawi | 41 | 136 | 43 | 44 | 82 | 112 | 458 |
| The Gambia | 13 | 224 | 96 | 112 | 102 | 112 | 659 |
| Kenya | 32 | 82 | 55 | 25 | 51 | 9 | 254 |

**Table 4:**
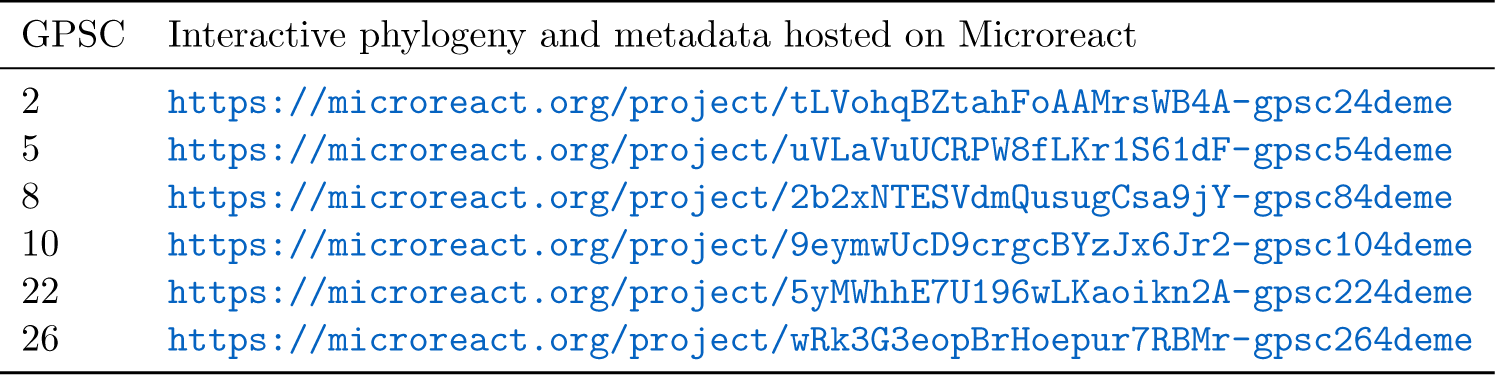
Interactive phylogenetic trees for each GPSC presented in the web server Microreact GPSC Interactive phylogeny and metadata hosted on Microreact.

**Table 5:** Country summary for between-deme migration analysis. Description of the four countries between which we estimated migration including number from each country, years of vaccine introduction and dosing schedule, the proportion of the dataset for each country which comprised NVTs and the percent of isolates from each country which were from children *<*5.

| Country | N | Vaccine Introduction | Dosing Schedule | %NVT | %Children <5 |
| --- | --- | --- | --- | --- | --- |
| South Africa | 6919 | PCV7: 2009<br>PCV13: 2011 | 2+1 | 31.91 | 65.2 |
| The Gambia | 3090 | PCV7: 2009<br>PCV13: 2011 | 3+0 | 56.70 | 46.4 |
| Malawi | 1612 | PCV13: 2011 | 3+0 | 42.68 | 49.6 |
| Kenya | 961 | PCV10: 2011 | 3+0 | 53.28 | 80.3 |

We included 81 ‘neutral’ non-antibody binding type genes selected from the pangenome wide immunological screen conducted by Croucher *et al*. [2017] Croucher et al. [2017]. These genes were core (minimum 99% prevalence) across each of the Dominant GPSCs. Further, we excluded linked SNP sites with a greater than 0.5 *r*^2^ relatedness threshold. We explored the Jaccard and Hamming distances between genomes for both the 81 and 355 genes and found clear boundaries between the country clusters with Hamming distances ranging from 0-0.5. Some GPSCs had clear distinguishable divergence between specific countries while other country pairs were very similar by Hamming distance (Figure S7, Figure S8).

### 3.3 Estimating the Weight and Direction of Migration for Country Pairs

To determine the symmetry of migration across all four demes included in this model (South Africa, The Gambia, Kenya, and Malawi) we considered 6 separate models, one for each GPSC, estimating the migration parameters between deme pairs. We used a uniform prior with bounds corresponding to the sensitivity of the simulation demonstrated in the *F_st_* sensitivity analysis (Figure 1A).

We implemented the two deme model to estimate migration parameters between each pair of demes asymmetrically given the 81 concatenated neutral genes with linked sites removed. We fit the model using BOLFI approximation and drew a posterior sample from it. We ran 3 independent implementations. We successfully estimated 10 of the 12 asymmetric migration parameters for all deme pairs by GPSC (Figure S13, Figure S14, Table S1). Parameter estimates for GPSC10 and GPSC26 between South Africa and Kenya did not settle on a single migration parameter in either migration direction due to the bimodal distribution of the posteriors and a co-linear relationship between the parameters. A migration weight which was higher from South Africa to Kenya than from Kenya to South Africa resulted in a similar model as the reverse; as such the model was unable to resolve these parameters (Figure S14). Resolving the peak which is ‘best’ is likely futile in that both peaks are equally likely to be true (Figure S14).

To summarize and compare migration between demes across GPSCs We divided the median posterior value by the maximum bound to find the probability of migration for each parameter. We then found the distribution of migration probabilities for each deme where the confidence intervals represent the distribution of values across GPSCs (Figure 3A). The difference between the estimates for each deme pair is the relative probability of migration. For example, the pneumococcal populations of South Africa and Malawi are consistently better explained by 50% more migration from South Africa to Malawi than the reverse, while the distribution between Malawi and The Gambia is explained differently across GPSCs with on average more migration from Malawi to The Gambia. The exception to this deme pair is in GPSC2 where the reverse is true (more migration from The Gambia to Malawi) (Figure 3B, Table S1).

**Figure 3:**
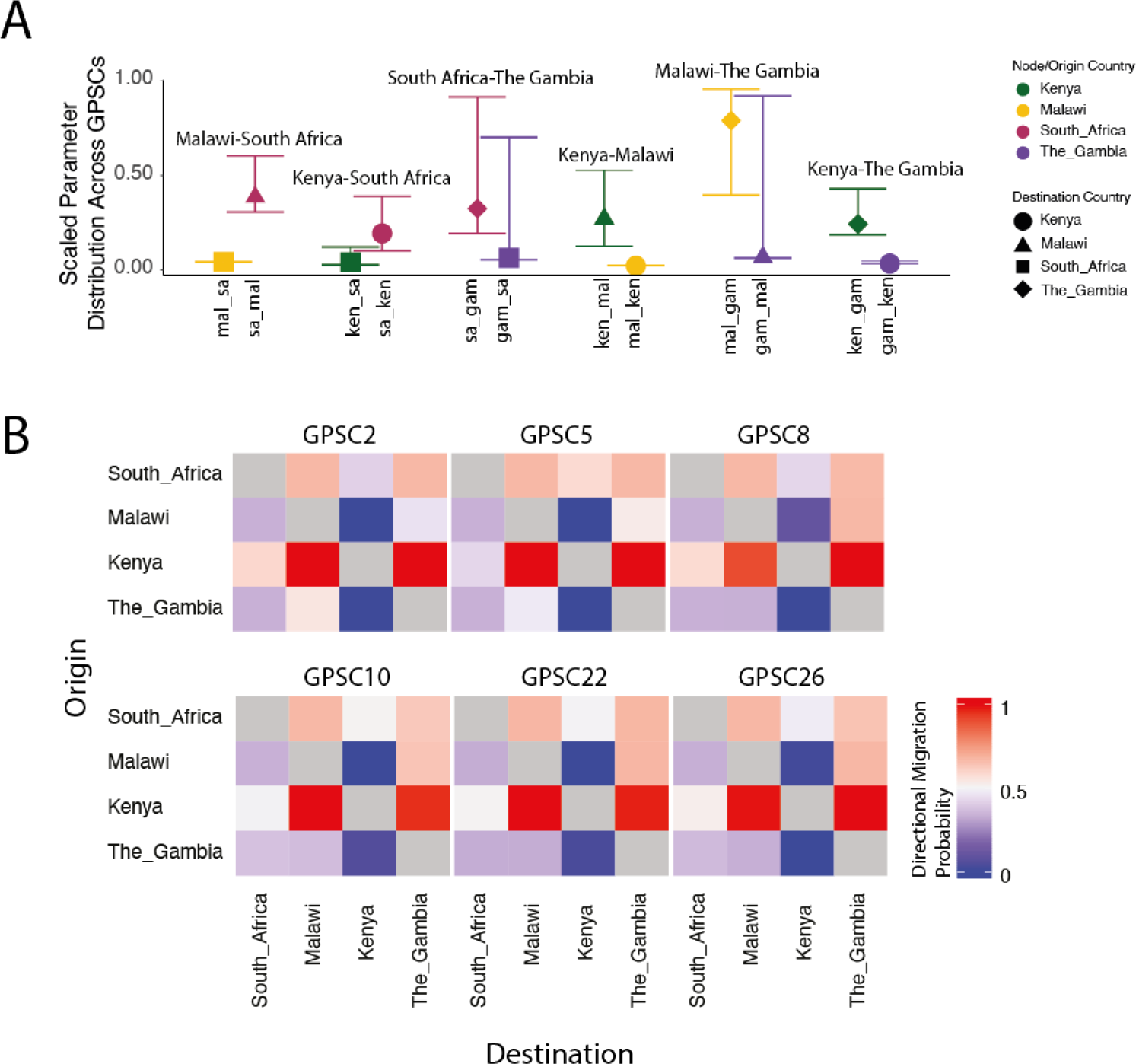
Migration parameters summary estimates. A)The scaled distribution of migration probabilities for each deme across GPSCs grouped by deme pair and colored according to country of origin and shaped according to destination. B) Migration parameters directional probability estimates between each deme pair. Colored by the probability of migration between each deme pair. where red is *>*50% migration probability and blue is *<*50% migration probability.

#### 3.3.1 Weight

To normalize the migration parameter estimates for each GPSC We found the mean parameter estimate across all posterior samples for each GPSC (*mig_gpsc_*). We then calculated the relative migration parameter estimate by dividing each parameter by the GPSC specific parameter 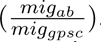. We first group these by deme to determine if there are consistent deme-wise patterns(Figure S15A) and then group them by GPSC to identify the differences across demes (Figure S15B, Table S1).

#### 3.3.2 Direction

We estimated the probability of directional migration for each GPSC and deme pair by identifying the percentage of posterior migration parameter estimates for *mig_ab_* which were greater than those for *mig_ba_*. We then grouped these into high (≥ 0.6), medium (0.4 − 0.6), or low (≤ 0.4) migration probability. There were two migration patterns. Pattern one is seen in GPSC26, GPSC22, GPSC10, and GPSC8 in which there was *only* symmetric migration between Kenya and South Africa and asymmetric between all other deme pairs. (Figure 3B). Pattern two applied to GPSC5 and GPSC2 and included symmetric migration between Kenya and South Africa as well as between Malawi and The Gambia, where all other pairs had asymmetric migration patterns (Figure 3B). Across all GPSCs there was a higher migration probability from Kenya to Malawi and Kenya to The Gambia than from either of those to Kenya; and a higher migration probability from South Africa to Malawi and South Africa to The Gambia than from either of those to South Africa. Considering South Africa and Kenya have the highest population sizes (60.04 Million and 54.99 Million respectively in 2019) this implies that the higher the population size, its relative contribution to between pair migration is likely to be higher (Table 1). For those GPSCs with migration pattern one we estimated a higher migration probability from Malawi to The Gambia, again in line with this hypothesis (Table S1). The consistent directional patterns between GPSCs from independent models is reassuring and helps to validate our framework.

#### 3.3.3 Demographic Contribution to Migration

Using the raw parameter estimates from all two-deme asymmetric models we interrogated whether the origin population size (*α*), the destination population size (*β*), or the distance between deme pairs at the centroid (km) (*γ*), had a larger contribution to the migration parameter estimates (*θ*). We fitted four logistic models, one for each parameter, and one model including all three parameters (Equation 5, Equation 6, Equation 7, Equation 8). In the model encompassing all three parameters a greater destination population size was significantly associated with the parameter estimate (*p* = 1.07*e^−^*^06^), while the origin population size (*p* = 0.606), and distance (*p* = 0.877) were not associated (AIC=150.13)(Equation 5).

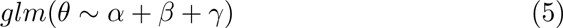

In both the overall model (Equation 5) and the individual model (Equation 7) a greater destination population size was again significantly associated with a smaller migration parameter (Overall: Coefficient −0.021, *p* = 1.07*e^−^*^06^, AIC=150.13; Individual: Coefficient=-0.021,*p* = 1.02*e^−^*^08^, AIC=146.66). The distance between countries was marginally associated (Coefficient 1.014e-04, *p* = 0.0354; AIC=176.05), but surprisingly in that a larger distance resulted in a higher migration parameter. This is largely driven by GPSC26 where the highest migration parameters are associated with The Gambia which is the most distant country from Kenya, Malawi, and South Africa. Due to the low sample size and limited distances explored the association between migration and distance is not robust or generalisable. In none of the models was the population size of the origin significantly associated with the migration parameter estimate (*p* = 0.232; AIC=179.16)(Figure S18). To be clear this applies specifically to these four countries and is an oversimplification of reality due to the discrete number of distances between the four countries explored in these migration models. However, in summary, the raw migration parameter estimates are negatively correlated with an increasing destination population size. This is sensible when placed alongside directional probability estimates whereby the higher population size is more likely to be the source of migration. Taken together for these GPSCs migrating between South Africa, Malawi, The Gambia, and Kenya this implies that the most migration is from large origin population sizes to smaller destinations.

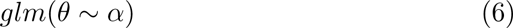

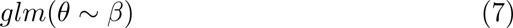

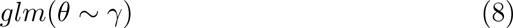

### 3.4 Estimating the Weight of Migration Across Four Countries

#### 3.4.1 Overview

We estimated symmetric migration parameters between four demes (6 migration parameters)(Figure S16). We expanded our prior to estimate parameters between the bound of 0 and 5 in line with the prior predictive analysis (Figure S3).

#### 3.4.2 Weights

To normalize the migration parameter estimates within each GPSCs we repeated the same method described in Section ‘Weight’. For GPSC2 and GPSC5 migration between The Gambia and Malawi exceeded all other deme pairs in line with what was seen in the 2 deme model. For GPSC8 the same pattern extended however migration between South Africa and Kenya followed closely behind. For GPSC26 migration between Kenya and South Africa was dominant with migration between The Gambia and Malawi being second highest. For GPSC10 migration between South Africa and the Gambia exceeded all other pairs and for GPSC22 migration between Kenya and The Gambia, and South Africa and The Gambia were lowest while all other deme pairs were similar (Figure 4, Table S2). In a generalized linear model there is no association between the distance between demes (Equation 8). However, when comparing the relative population size between deme pairs to the migration parameter estimates there is a significant association (Coefficient=-0.027, *p* = 0.00324; AIC=101.13) in that as the relative population size increases between demes there is less migration between them.

**Figure 4:**
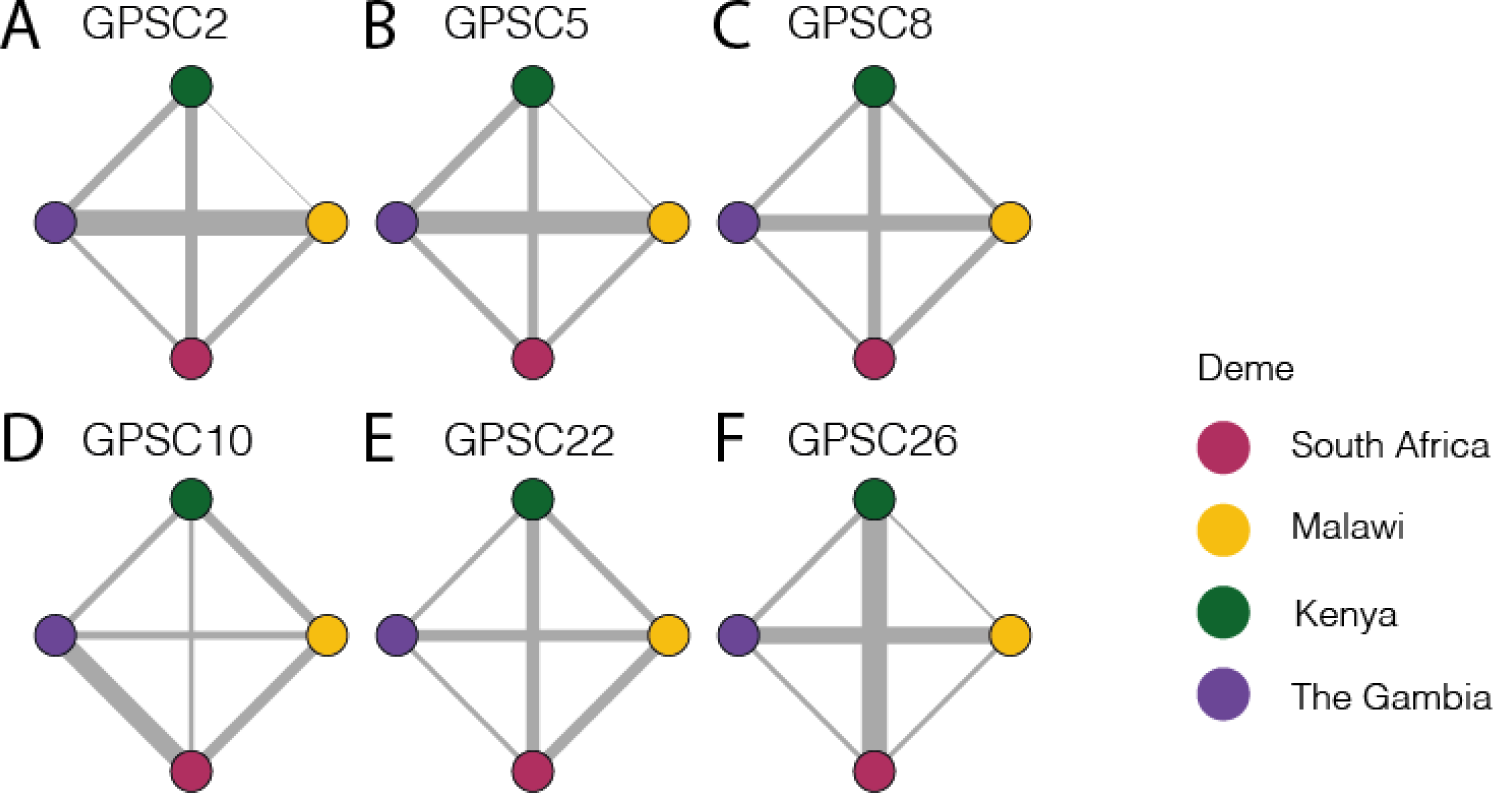
Migration parameter estimates for each GPSC. Migration paths weighted by the relative migration rate for each deme pair within each GPSC. The nodes are colored as follows: South Africa is represented in pink, Malawi in yellow, Kenya in Green, and The Gambia in purple.

### 3.5 Between-country migration probability

While here we are able to identify migration parameters when migration occurs we do not account for how often migration may occur but within the same country, or not occur at all. To address this we used a risk ratio framework to investigate the risk of similarity across geographic distance Belman et al. [2023], Salje et al. [2017], Lefrancq et al. [2022]. We found that after 43 years of spread pairs are equally likely to be in Kenya as between Kenya and another country with an RR of 2.55 (95%CIs 0.42-8.19), for South Africa this is 55 years with a RR=1.55 (95% CIs 0.75-6.63), for The Gambia it is after 53 years of spread; RR=1.82 (95% CIs 0.82-3.36), and for Malawi this is after 55 years of spread; RR=2.20 (95%CIs 0.35-4.92)(Figure 5).

**Figure 5:**
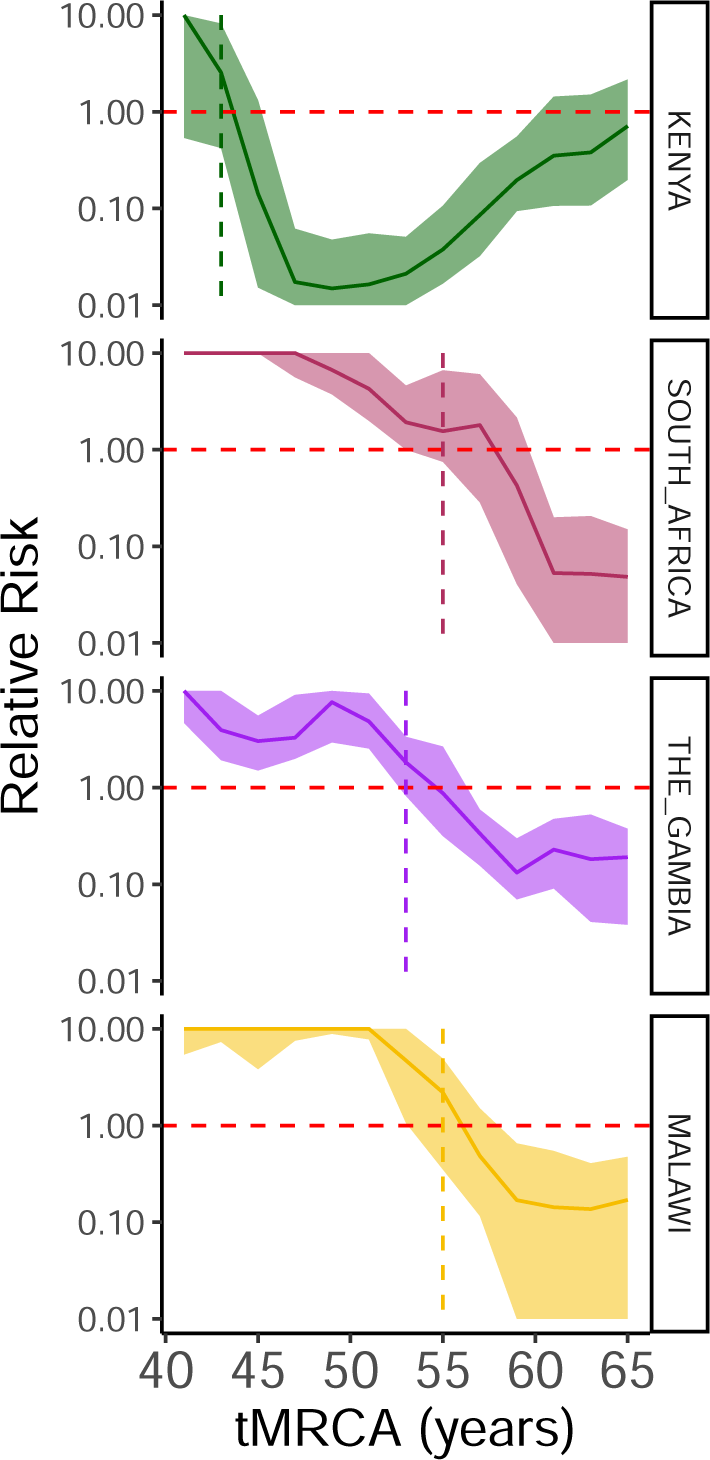
Out-of-country migration probability. The risk of pairs within divergence times across rolling 10-year time windows of being found within the same country as between each country and every other country for Kenya (green), South Africa (pink), The Gambia (purple), and Malawi (yellow). A dashed red line indicates a relative risk=1 while a vertical dashed line for each plot denotes the tMRCA at which the relative risk crosses 1.

### 3.6 Discussion

Our modeling framework utilizes genomic data and population summary statistics to infer migration parameters between demes that can best replicate the extant population distribution. We were able to successfully validate the method using simulated data both estimating the direction of migration between demes in the two-deme model, as well as the weight of migration across four demes. Interestingly, we detected some heterogeneity in migration rates across different lineages. The baseline expectation is that considerable variation would not exist between GPSCs. The similarity in migration between invasive and non-invasive lineages may imply, as found in Tonkin-Hill *et al*. Tonkin-Hill et al. [2022] that although the invasive lineages are not frequently found in carriage, they may persist at low population frequencies allowing them to be transmitted across borders in similar patterns (Figure S17).

We also find country-pair specific migration patterns which are in line with the source country usually being that with the greater population size when inferring directionality. However, when determining the demographic factor (population size of origin or destination and distance between them) with the greatest contribution to migration we found that the destination population size was more important.

Some limitations to this method include our inability to include the true carriage population sizes and true genome length within the simulation framework. As such, it is impossible to contextualize the migration parameters within the context of time, however, their relative relationship remains useful nevertheless. Further, as we use the population sizes for each deme in accordance with the true population sizes of those countries, some bias may be introduced since the pneumococcal carriage rate is known to vary by both country and human population. The age structure of each deme may consequently influence the true carriage rate in that a deme with a larger child population relative to the adult population may have more carriage overall, while simultaneously, a population with more children is unlikely to be as mobile as an adult population. Resolving this would require more demographic interrogation of pneumococcal carriage and human mobility in these regions. Our framework could also be applied on a smaller spatial scale where more granular data exists about the underlying carriage rates, and migration between regions, provinces, or states could consequently be inferred with better precision. Additionally, the ability for the migrating bacteria to take hold in a country depends on previous pathogen spread as well as vaccine campaign implementation.

Future implementations of our models could for example include parameter estimates for the population size of each deme, and incorporation of vaccine coverage and future immunity. Alternatively, the migration parameters could be incorporated into independent migration simulation frameworks which account for population immunity and other covariates. Human mobility data could in theory be used but currently (September 2023) representative between country human-mobility data remains sparse. Meta provides travel data between-countries but these are dominated by high income countries. Of the four demes included in our framework only South Africa is present within the Meta datasets from 01/2021-04/2023. Interrogating open flights (https://openflights.org/data.html) between-country data for these four countries only provides sparse data for Kenya and Malawi, and no data for The Gambia. The majority of flights within Africa are not taken directly, they often include many stopovers which would further complicate the use of such data. Further, flight data would only represent a small subset of possible movements and in low-income countries it is largely reflecting tourism, while not accounting for the movement of the majority of the population Gössling and Humpe [2020], Findlater and Bogoch [2018].

Estimates of migration rates such as those produced as part of this study could be informative for vaccine implementation policy. If there is more migration from deme A to B, implementing the vaccine first in deme A may have spillover effects into deme B, allowing such interventions to have an effect beyond country borders. Some possibilities for further development of our framework include incorporating additional countries, and inferring correlation between GPSC-specific migration patterns and classes of mobility such as flights compared to roads; or adults compared to children. This will help us to understand what types of mobility best explain the estimated migration parameters. Provided rich data from genomics-based surveillance systems, the current approach would also be applicable to multiple other species of bacteria.

## 4 Data availability

The code and data associated with this paper are available at https://github.com/sophbel/LFI_between_country_migration/tree/main.

## Acknowledgments

This work was supported by Wellcome under grant reference 108413/A/15/D, 2016194, and WT098051. For the purpose of Open Access, the author has applied a CC BY public copyright license to any Author Accepted Manuscript version arising from this submission. Further we would like to thank all Global Pneumococcal Sequencing Project partners particularly those at the National Institute for Communicable Diseases in South Africa, the Medical Research Council in The Gambia, the Malawi Liverpool Wellcome Trust, and the Kenya Medical Research Institute.

## 5 Funding

This GPS project was funded by the Bill and Melinda Gates Foundation (grant code OPP1034556). S.B. and S.D.B. were supported by the Wellcome Sanger Institute (core Wellcome grants WT098051, 206194, and 108413/A/15/D). N.J.C. was supported by the UK Medical Research Council and Department for International Development [MR/R015600/1, MR/T016434/1]; a Sir Henry Dale Fellowship, jointly funded by Wellcome and the Royal Society [104169/Z/14/A].

## 6 Conflicts of interest

N.J.C. has consulted for Antigen Discovery Inc and Pfizer, and been invited to attend meetings funded by M.S.D. N.J.C. has received an investigator-initiated award from GlaxoSmithKline. No other authors have conflicts of interest to report.

## A Supplementary Figures

**Figure S1:**
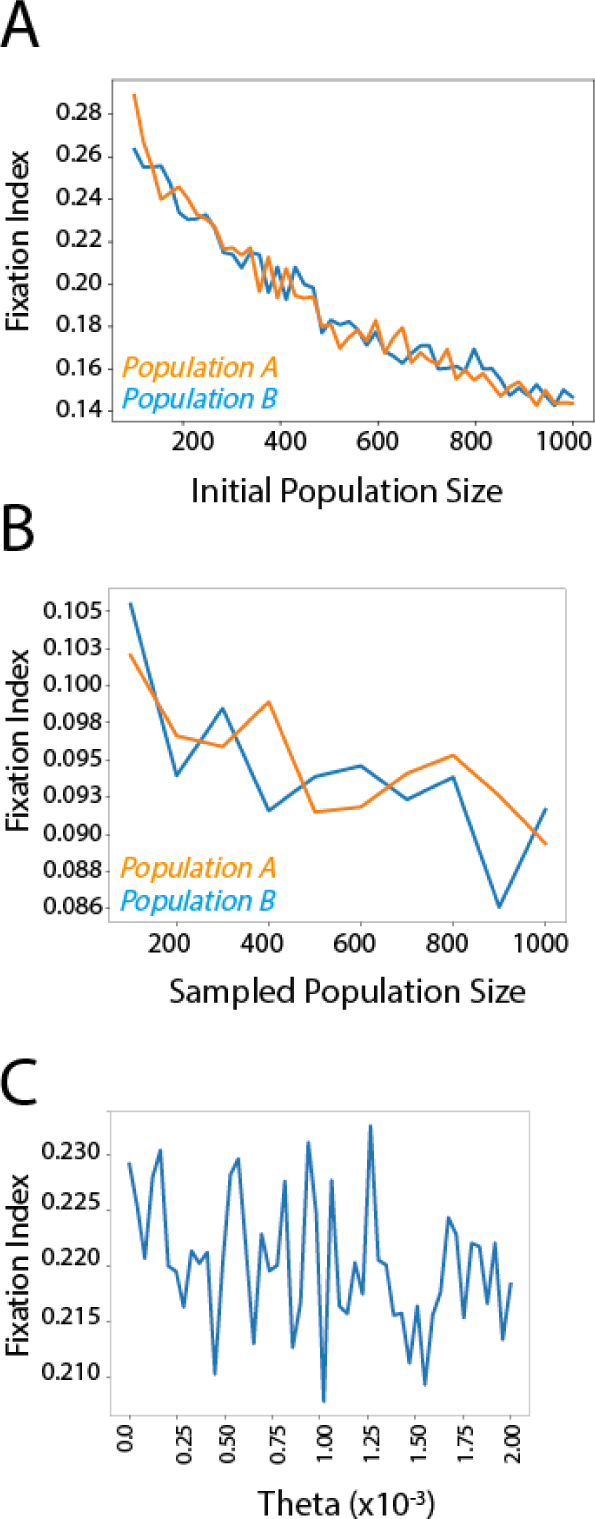
Sensitivity of the *F_st_* value to simulation parameters fixed at a sequence length of 100. A) Initial population size fixing the alternate population size at 500 and the migration parameter at 2, sampling 100 from each. B) Sampled population size with a fixed migration parameter of 2, the sample size for the alternate population fixed at 500 and initial population parameters for A and B at 600 and 200 respectively. C) *θ* parameter (mutation rate) with a fixed migration parameter of 2, initial population sizes of A and B 600 and 200 respectively, sampling 100.

**Figure S2:**
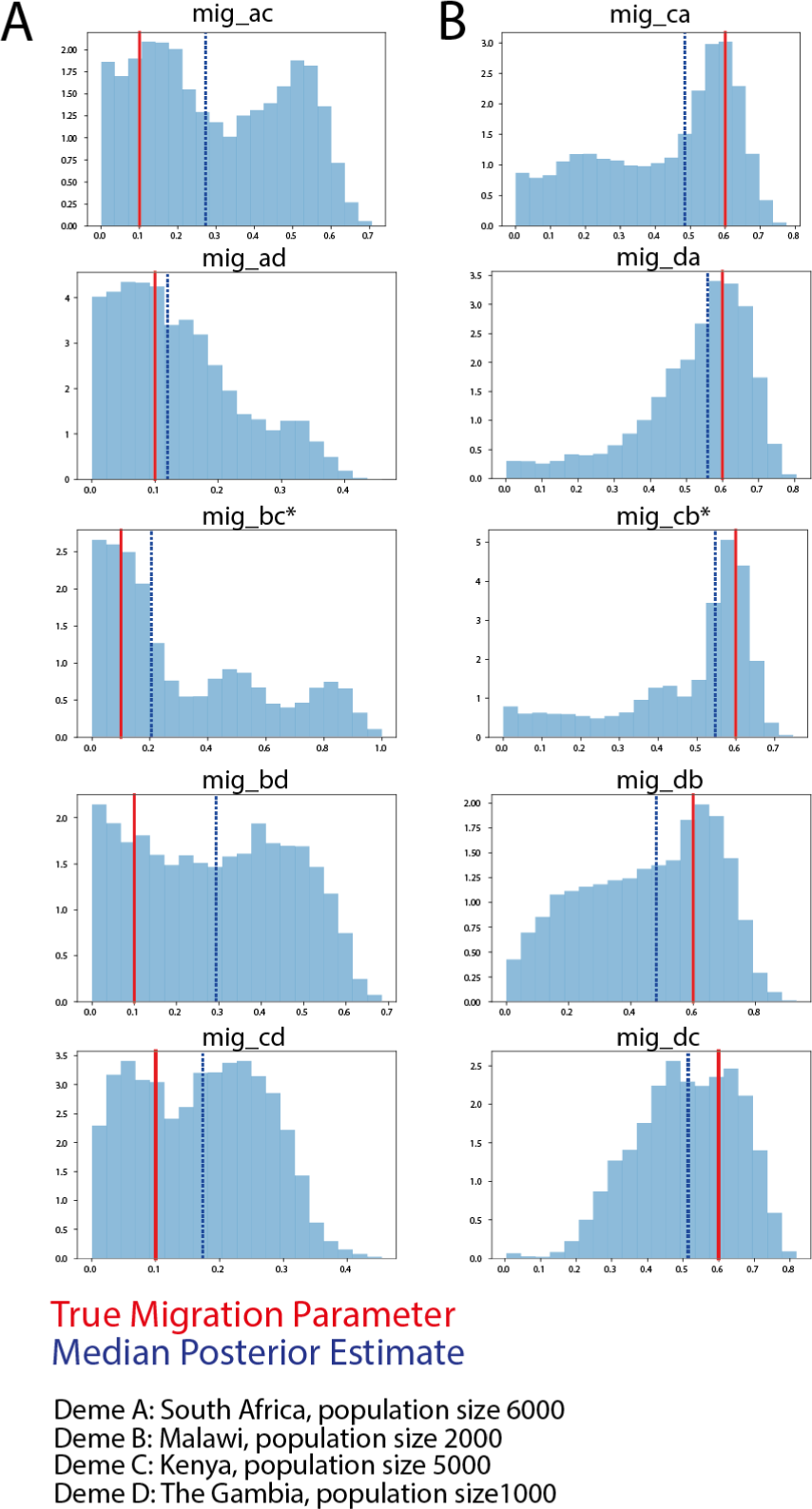
Recapturing input migration parameters with a 2 Deme model. A) The overlapping posterior density migration parameter estimates for migration parameter 1 — from a population [*a* − *d*] to population [*a*−*d*] B) and the inverse. The ‘true’ input parameters were *mig_a−d_*=0.1 and *mig_a−d_*=0.6. The posterior densities were estimated with a uniform prior and are visualized independently for each parameter (light blue), the true input parameter is indicated by the red vertical line while the median posterior estimate is indicated by the blue dashed line. Deme A=South Africa, initial population size 6000; Deme B=Malawi, initial population size 2000; Deme C=Kenya, initial population size 500; and Deme D=The Gambia, initial population size 1000. *Used no-uturn (nuts) sampling rather than metropolis sampling for *migbc* and *mig_c_b* due to difficulty converging

**Figure S3:**
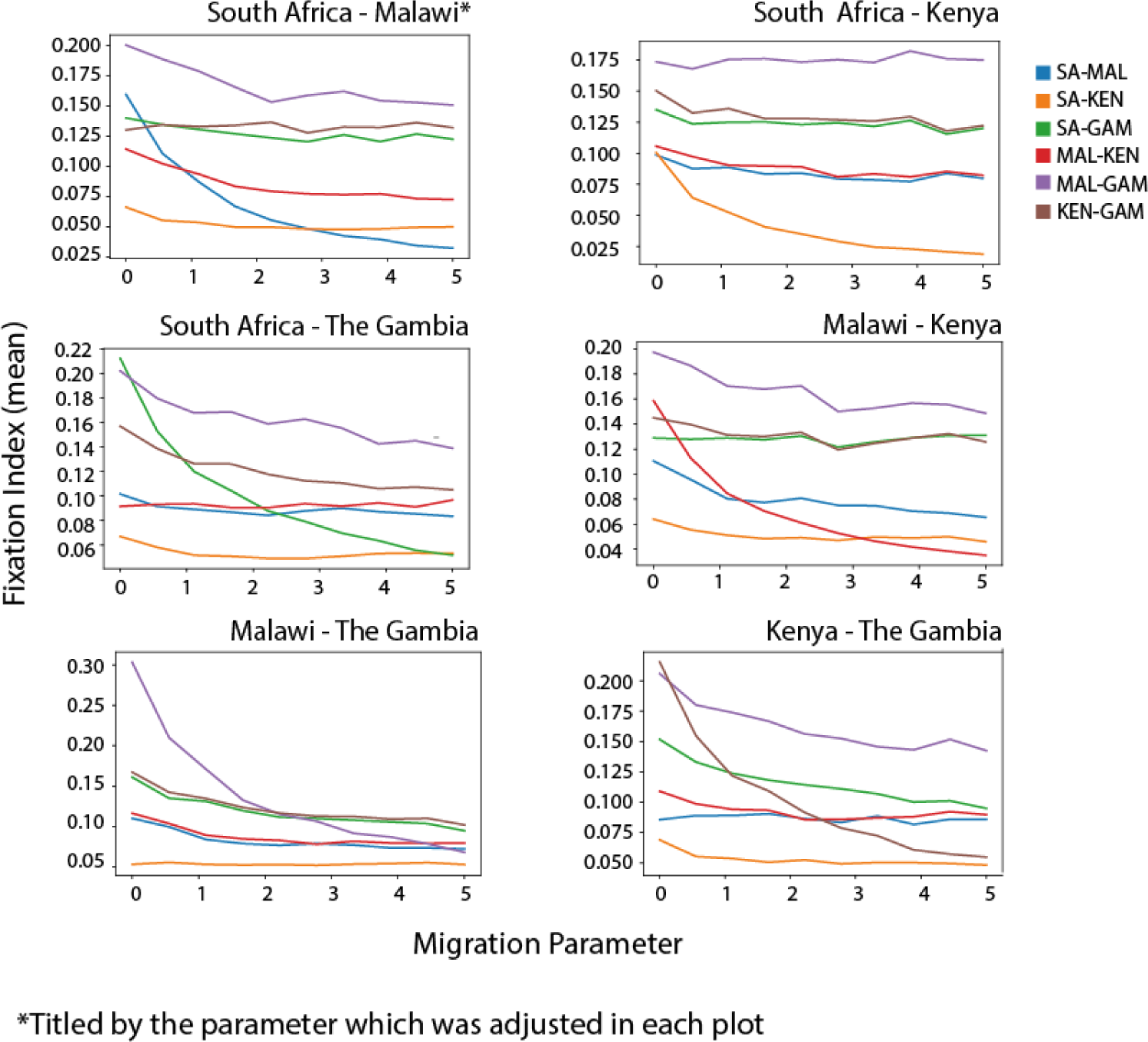
The response of the fixation index to varied migration parameters. Each plot indicates which migration parameter We varied and the *F_st_* between the countries for each of those migration parameters is indicated by the colored lines.

**Figure S4:**
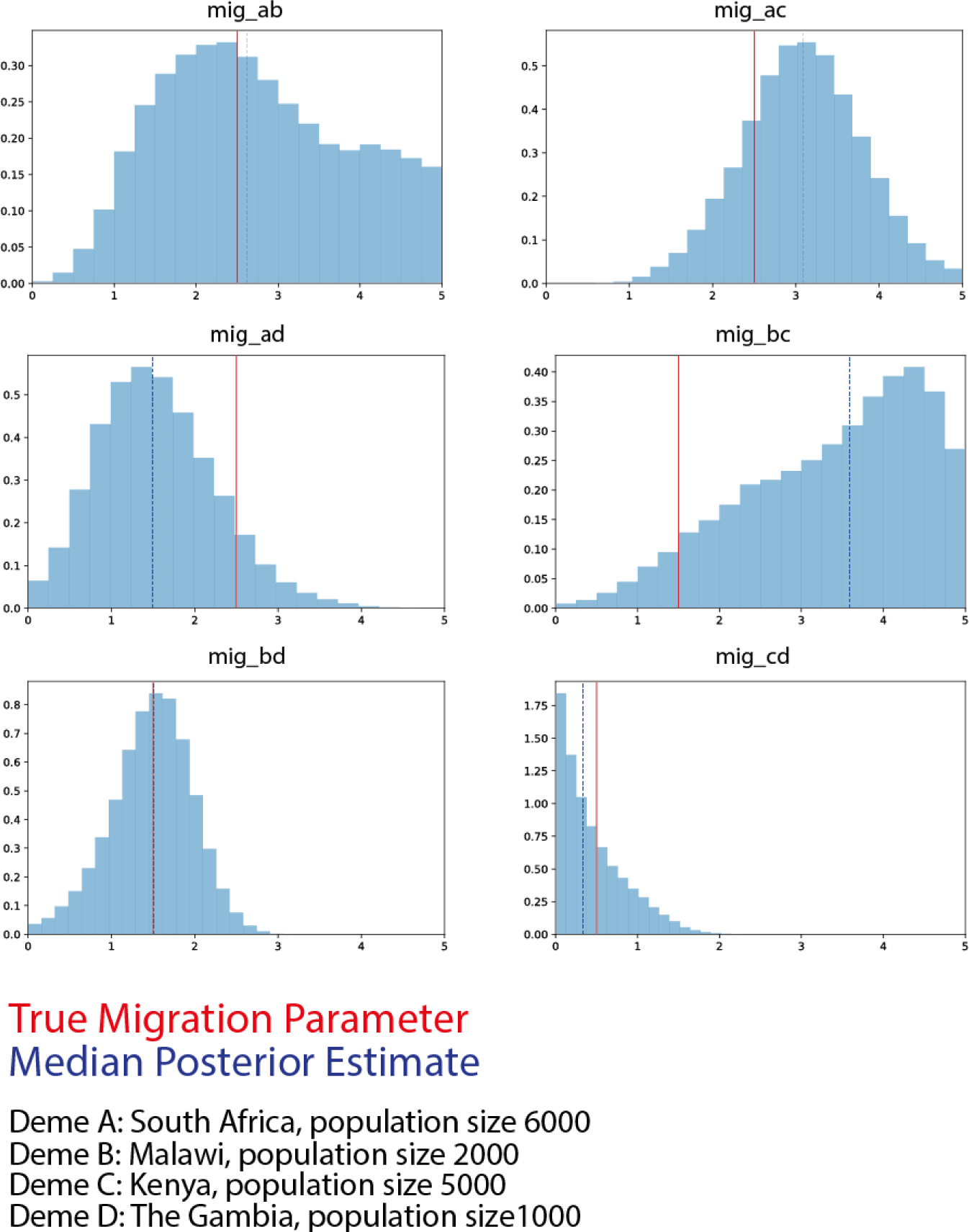
Recapturing migration parameters in the 4 deme model. The True Migration parameter is indicated by the red vertical line while the estimated median parameter is indicated by the blue-dashed vertical line. The posterior distribution density is represented by the blue histograms for each deme pair indicated by the title where a=South Africa, b=Malawi, c=Kenya, and d=The Gambia. The input population sizes for each of these scale to the true population size and are indicated in the figure.

**Figure S5:**
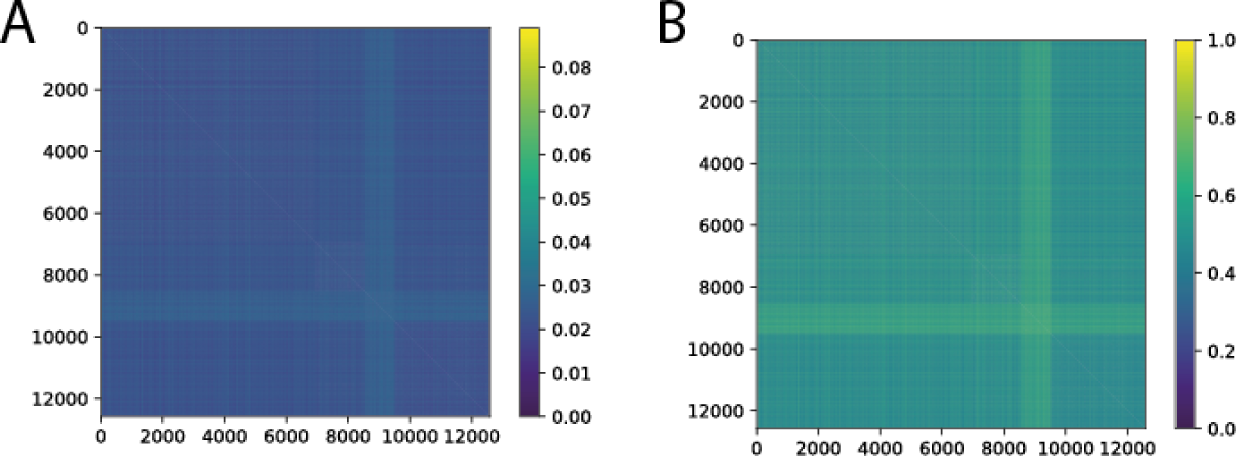
Pairwise distance estimates for between-country genomes across all 12,582 genome pairs from South Africa, Malawi, Kenya, and The Gambia, clustered in that order by A) Hamming distance and B) Jaccard Distance.

**Figure S6:**
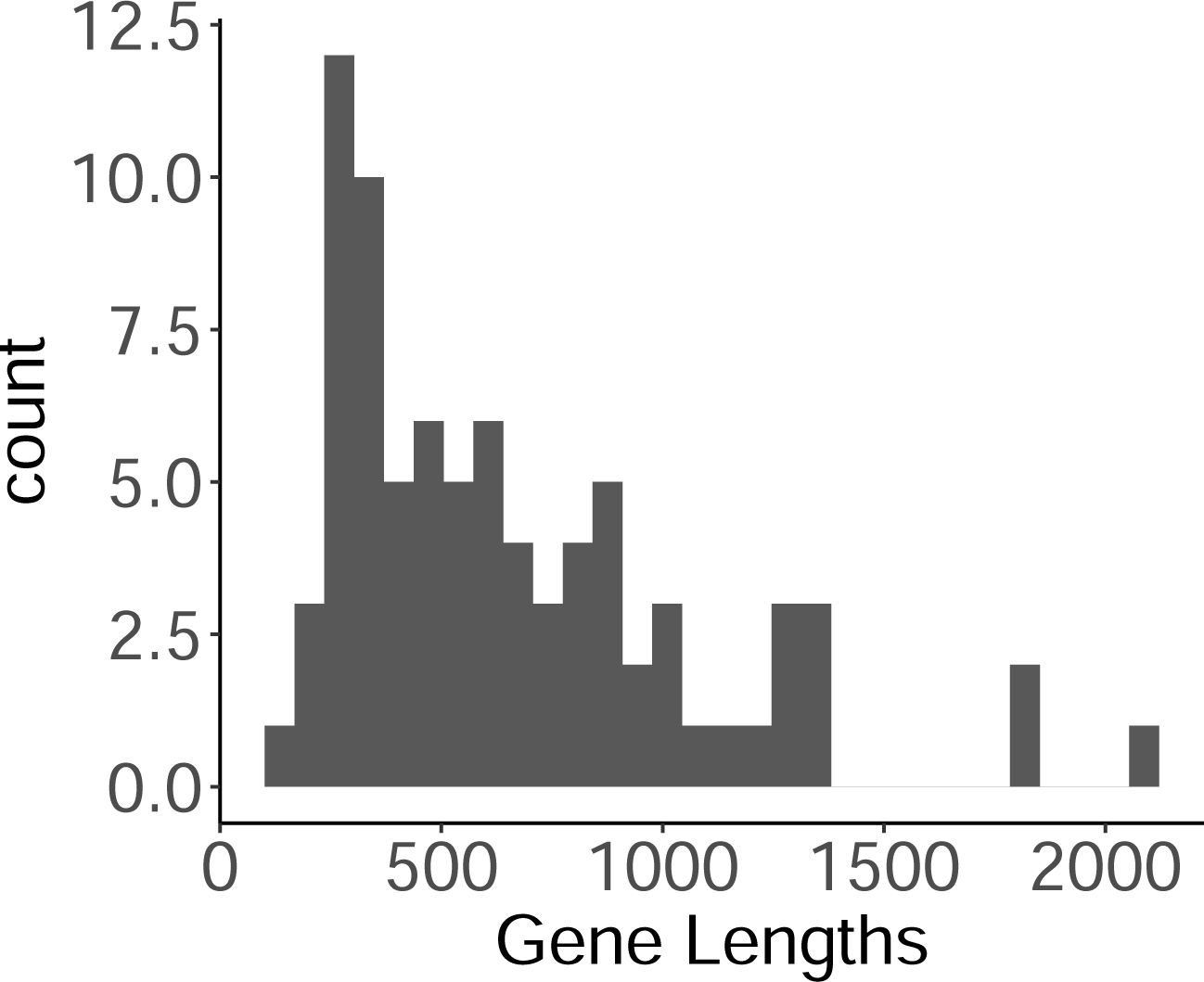
Histogram of gene length for each of the 81 neutral genes.

**Figure S7:**
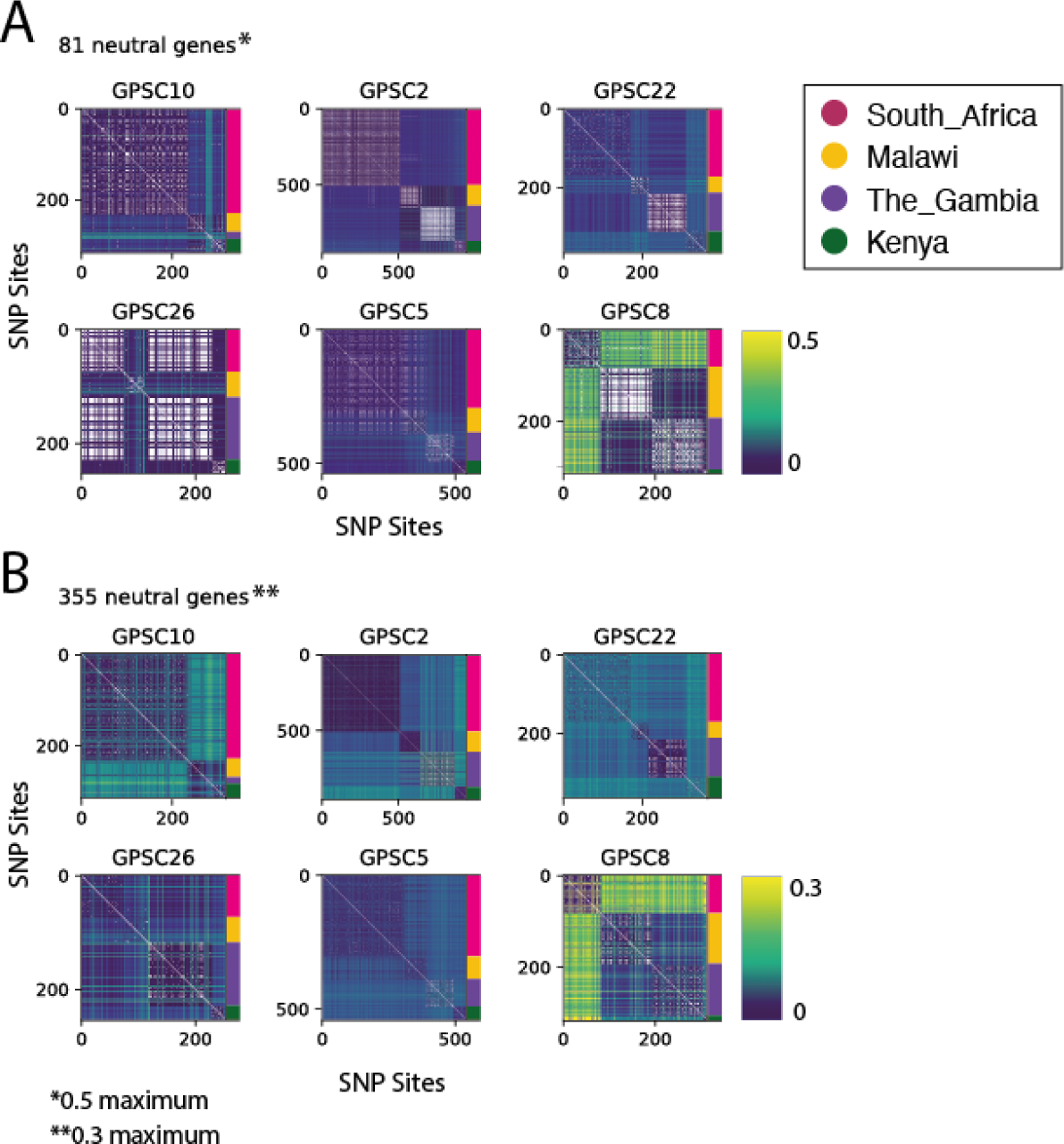
Pairwise Hamming distances across all genomes from each of the four demes (organized in the order of South Africa, Malawi, The Gambia, Kenya) for each GPSC in turn. These only include biallelic SNP sites. A) Includes 81 ‘neutral’ genes. B)Includes 355 ‘neutral’ genes.

**Figure S8:**
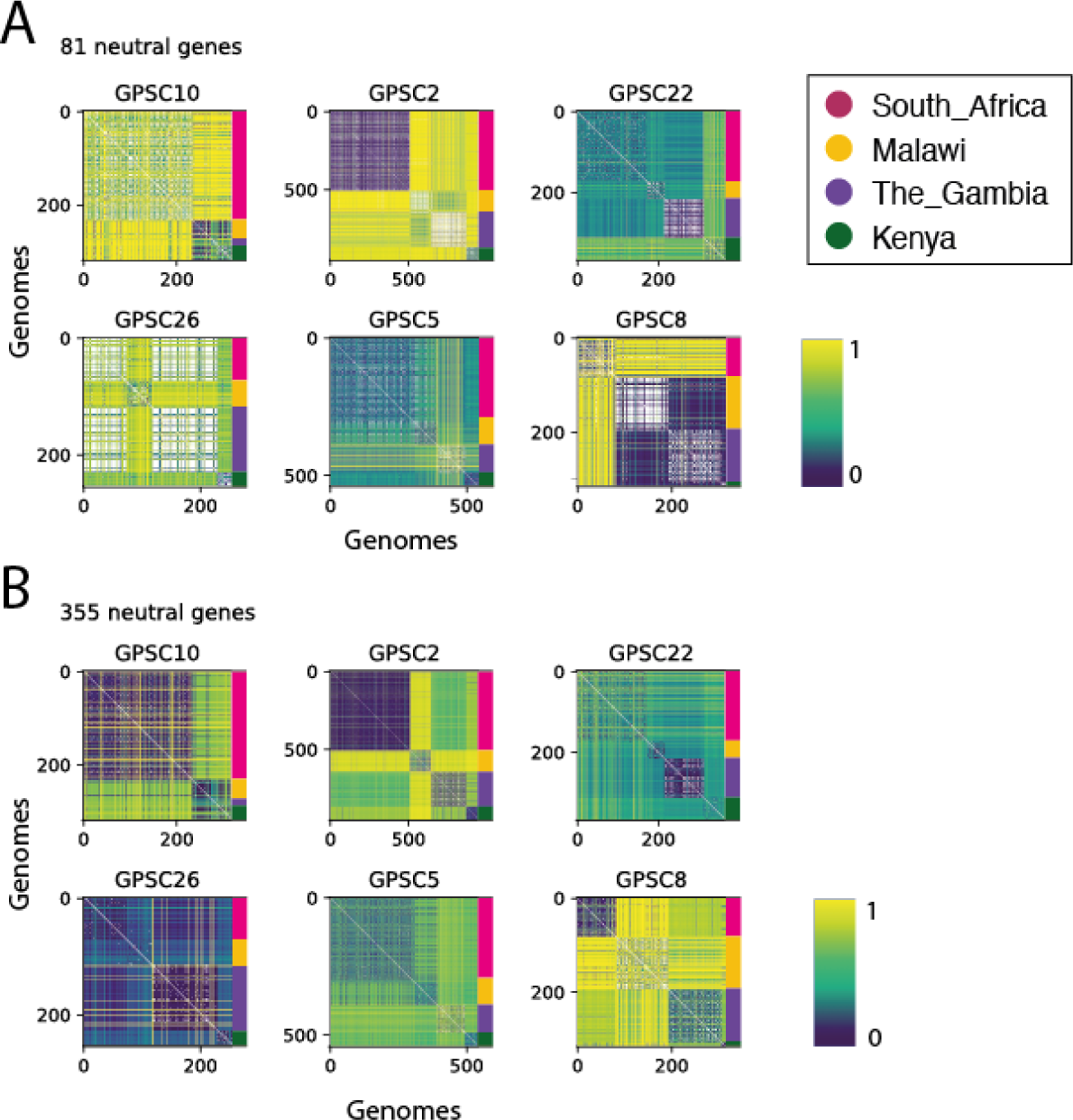
Pairwise Jaccard distances across all genomes from each of the four demes (organized in the order of South Africa, Malawi, The Gambia, Kenya) for each GPSC in turn. A) includes 81 ‘neutral’ genes, B) includes 355 ‘neutral’ genes. These only include biallelic SNP sites.

**Figure S9:**
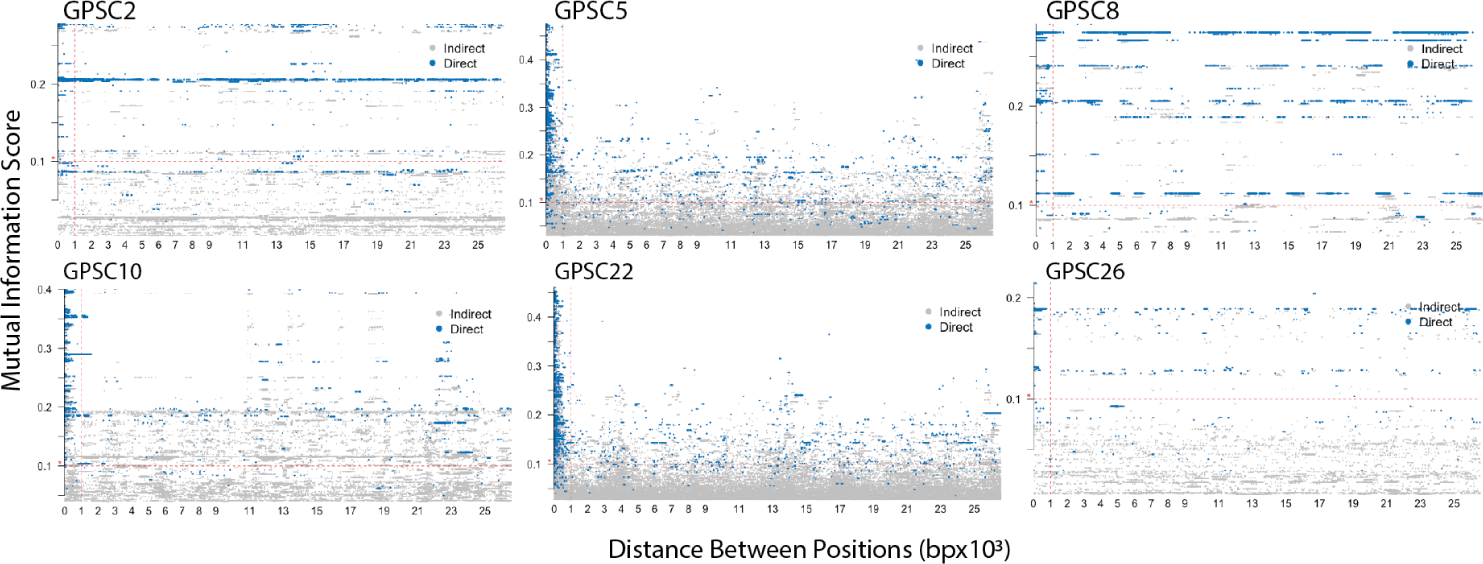
Mutual Information Scores between SNP pairs across 81 ‘neutral’ gene alignments for each of the GPSCs. The vertical dashed line indicates the 1kb cutoff under which removed correlated sites. The horizontal dashed line indicates the 0.2 mutual information score cutoff which has been used previously for the pneumococcus.

**Figure S10:**
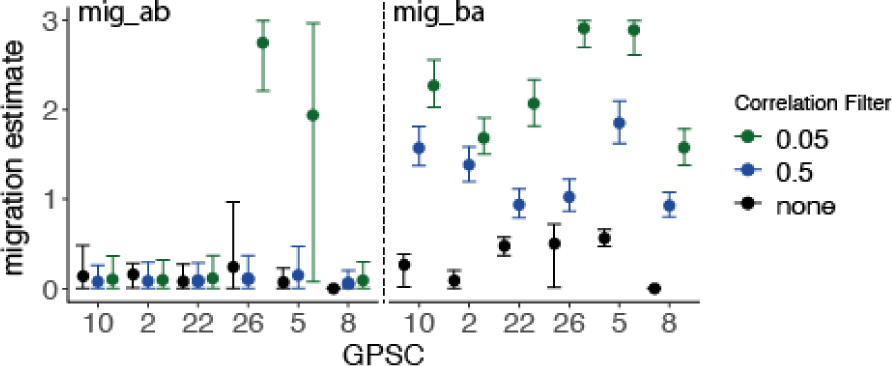
Estimated migration parameters removing correlated sites including *mig_ab_* on the left and *mig_ba_* on the right. Excluding all within a 1kb window upstream with an *r*^2^ *>* 0.5 (triangle), excluding all within a 1kb window upstream with an *r*^2^ *>* 0.05 (square), and retaining all sites (circle). The error bars indicate 95% CIs and each GPSC is along the x-axis. Initial population sizes were for South Africa (deme A) and Malawi (deme B).

**Figure S11:**
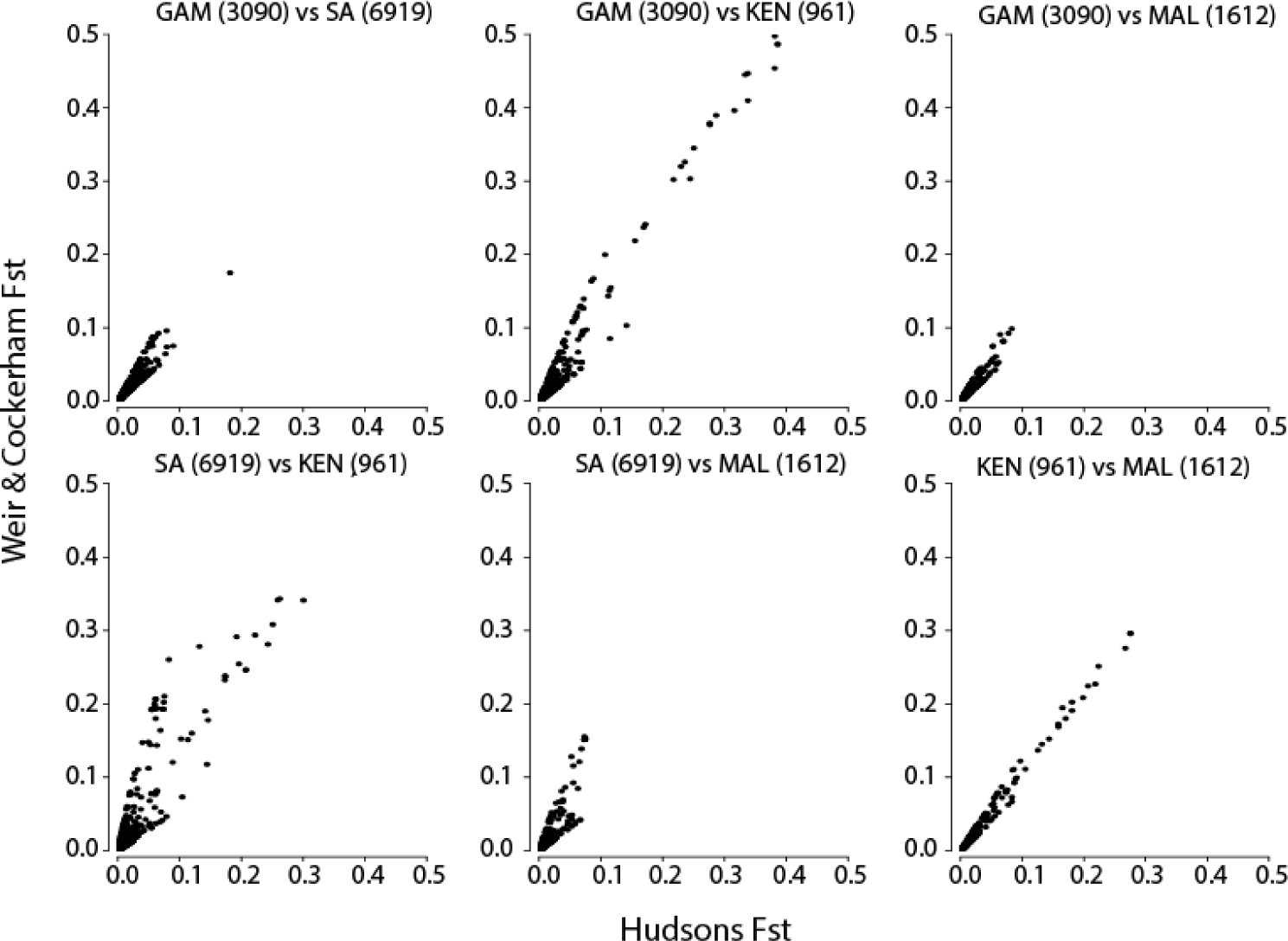
Pairwise comparison between the Hudson and Weir-Cockerham *F_st_* values across all four demes. In total this includes six comparisons, one between each deme and every other deme.

**Figure S12:**
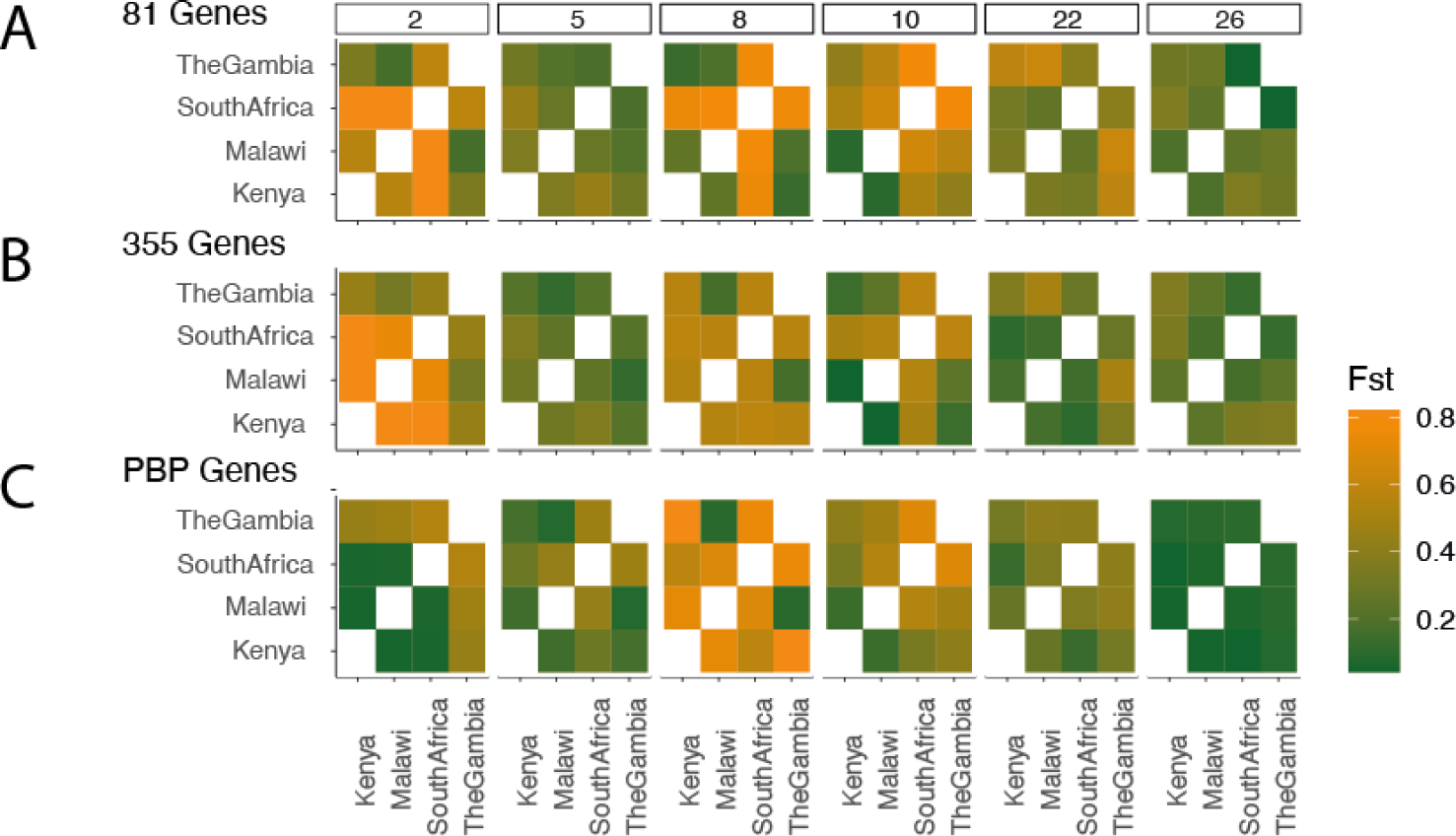
Hudsons *F_st_* across all genomes between each of the four demes for each GPSC. A) calculated from 81 genes, B) from 355 genes, and C) only including the PBP genes (which are likely under selection in each place due to their interaction with penicillin-resistance acquisition). A higher *F_st_* is a more divergent, separate population, while a lower *F_st_* is a more highly mixing population, also known as panmictic.

**Figure S13:**
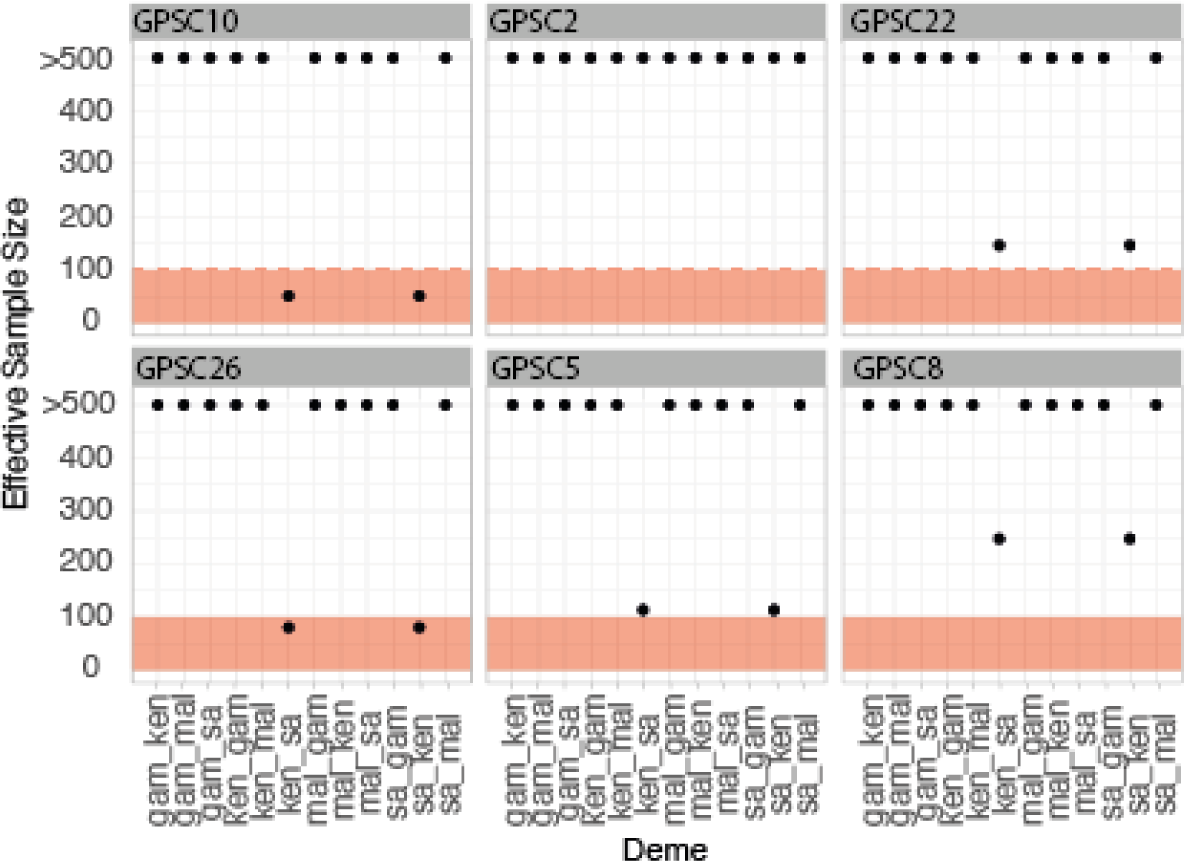
Convergence of asymmetric 2 deme parameter models. A) The effective sample size (ESS) across all parameters estimated. ESS *<*100 is indicated in red. B) The posterior density of parameter estimates between South Africa and Kenya for GPSC10. These were unable to converge due to the high co-linearity between them.

**Figure S14:**
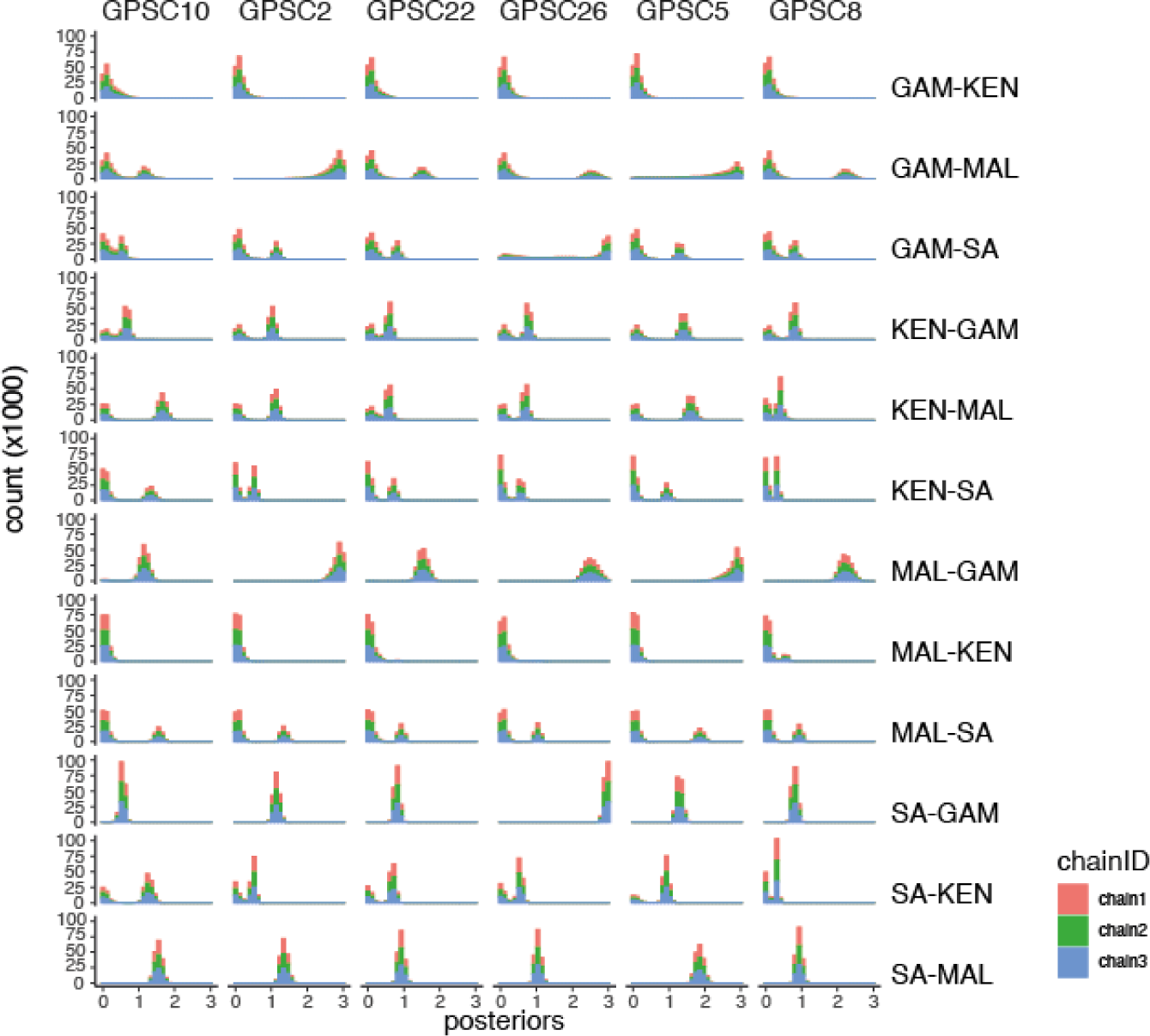
The posterior parameter distributions across 3 independent runs of 4 chains each across the 6 dominant GPSCs (columns) and 12 parameter estimates (rows).

**Figure S15:**
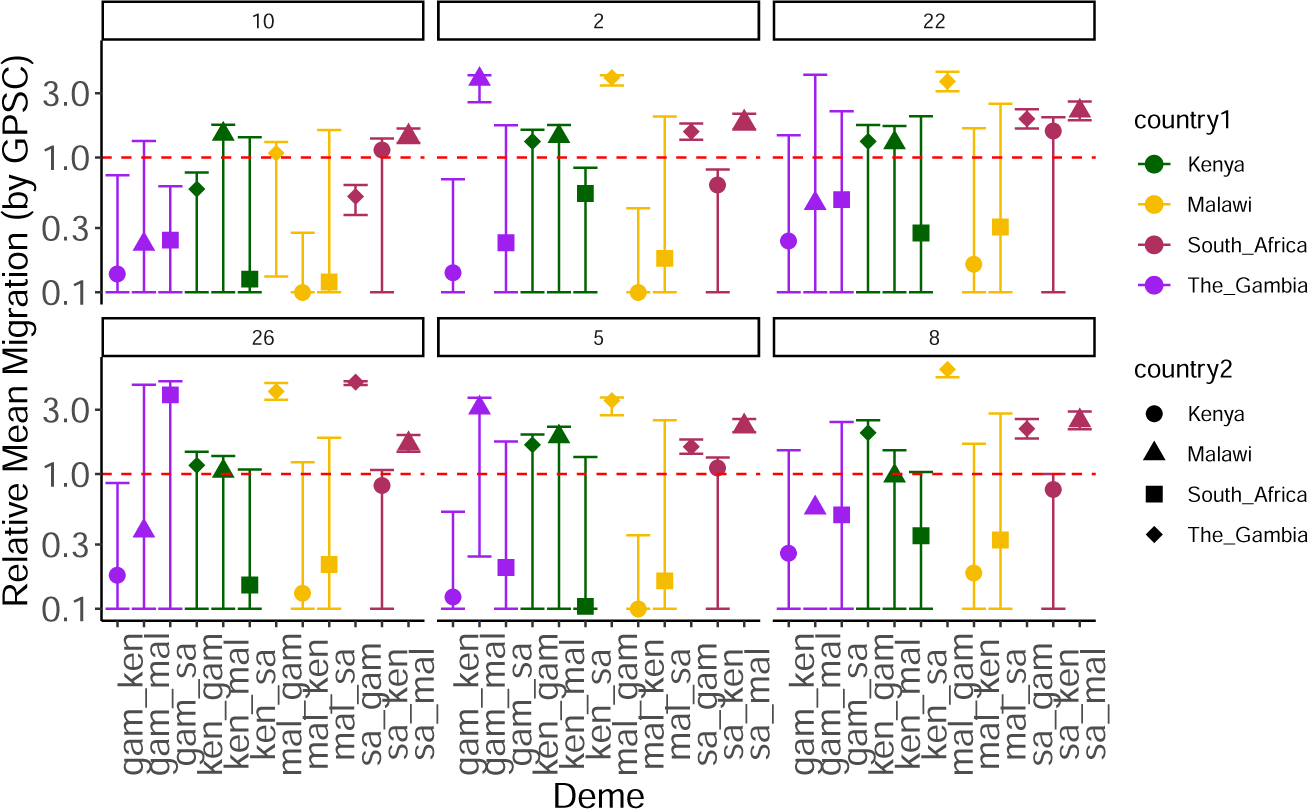
Relative migration parameters asymmetrically between two deme pairs. Relative migration for each deme pair within each GPSC independently. The x-axis indicated the deme and they are grouped by GPSC. The origin location of South Africa is represented in pink, Malawi in yellow, Kenya in Green, and The Gambia in purple.

**Figure S16:**
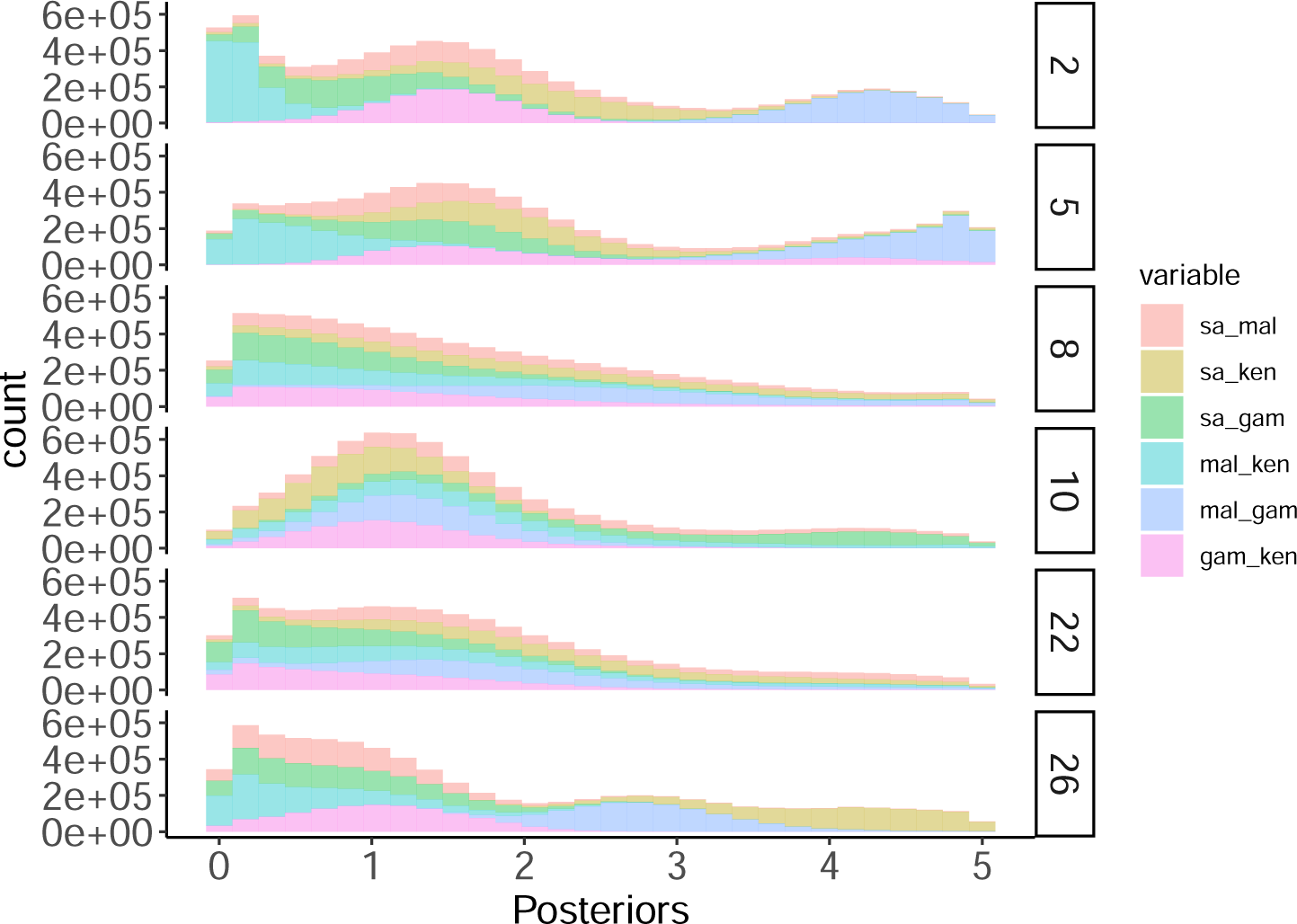
Posterior distributions for 6 parameter estimates for each GPSC, colored by parameter.

**Figure S17:**
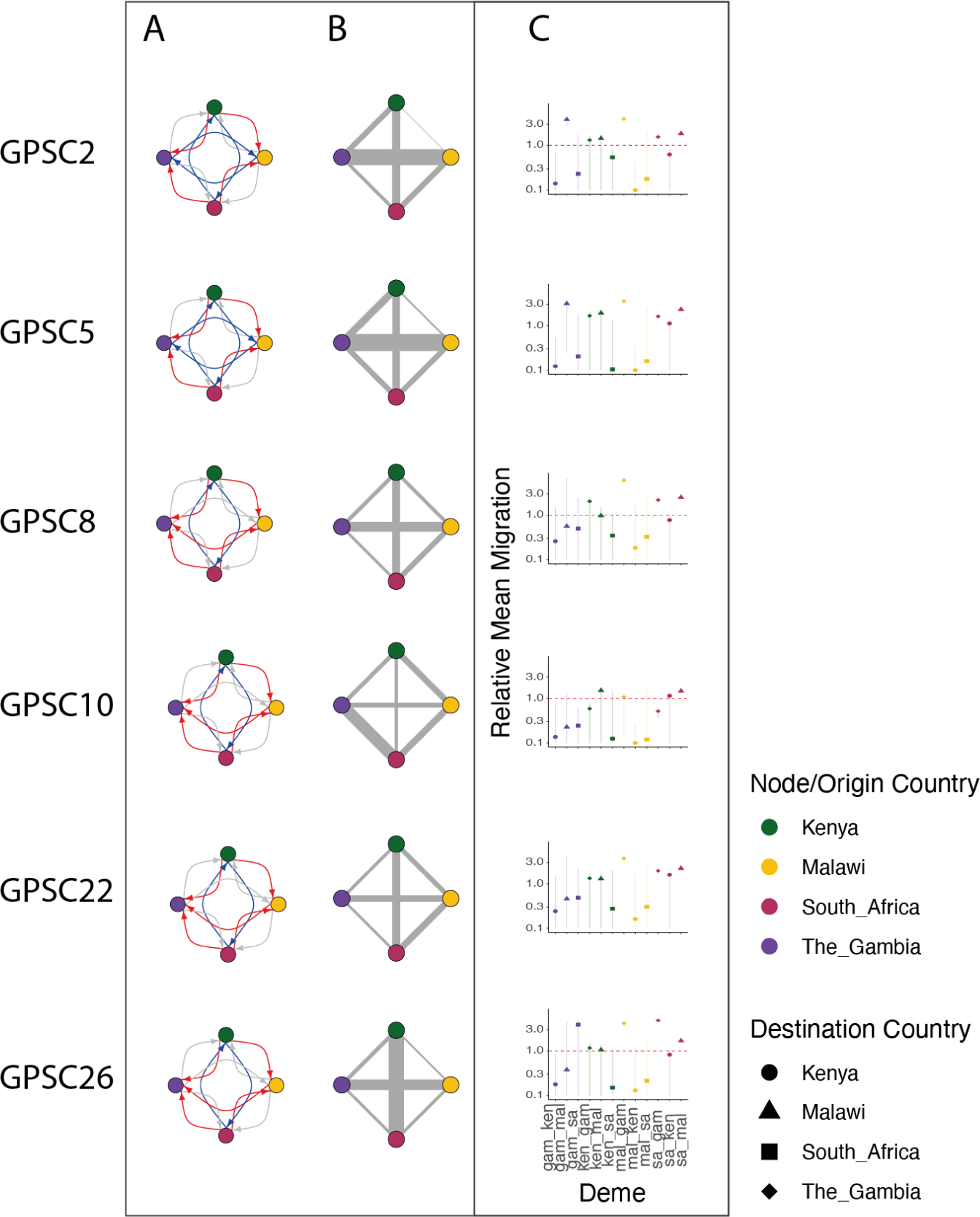
Summary of each GPSC migration parameters. A) The directional probability from the 2 deme model for each GPSC whereby red=*>*0.6, blue = 0.4-0.6, and grey = 0.1-0.4 probability of migration asymmetrically for each deme pair. The Node colors are described in the legend. B) The weighted migration from the 4 deme model between all 4 demes. The node colors are the same as A. C) The relative migration probability for each GPSC across all demes. The Origin country is colored the same as A and B and the Destination country is indicated in the legend.

**Figure S18:**
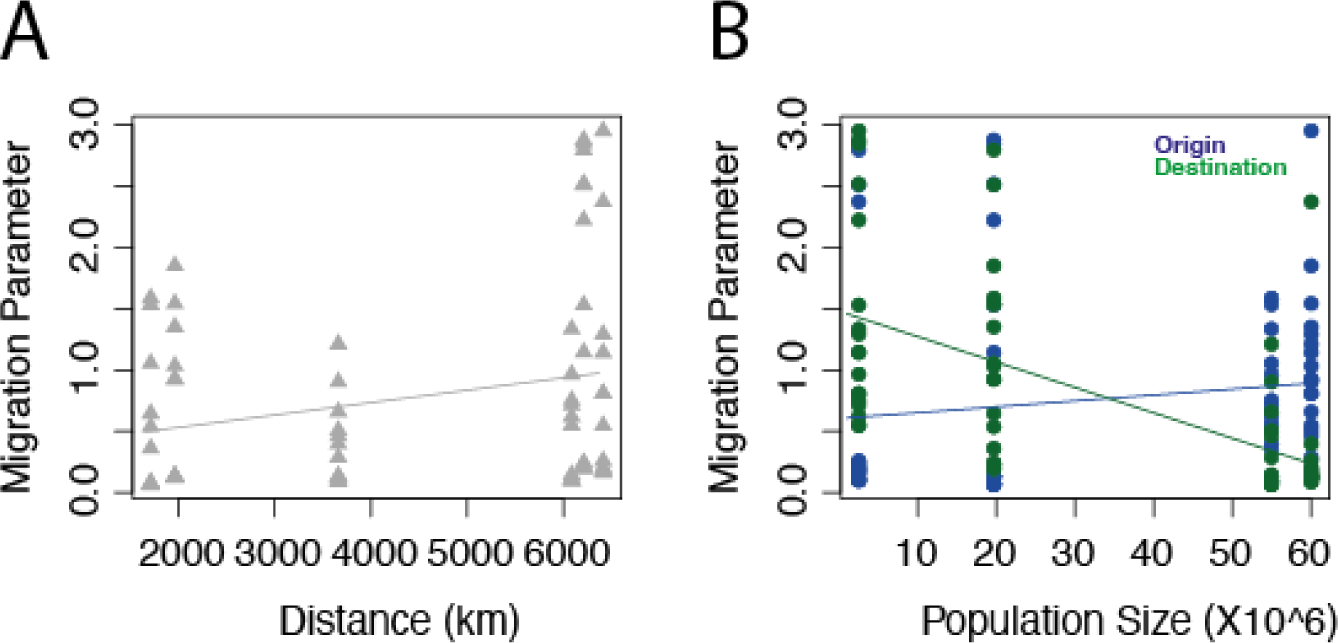
The population sizes and distance between countries versus migration parameter estimates. All plots include the migration parameter estimates (y-axis) against either distance between countries or the population size of the countries (x-axis). The left plot includes the distance between migration parameter demes (grey, triangles) and the right plot includes the population size of the origin (blue) or the destination (green). The models associated with each figure are included in lines of the same color.

## B Supplementary Tables

**Table S1:** Parameter estimates across all pairs within the two deme model. Values within the square brackets denote the 95% confidence intervals. The ‘Parameter’ is the raw migration parameter estimate while the ‘Relative Parameter’ is relative to all other deme pairs *within* each GPSC. The ‘Directional Migration Probability’ is the probability of migration asymmetrically for each GPSC and each deme pair (ie for *sa − mal*, GPSC10 there is 0.667 probability of migration while for *mal − sa* there is (1 *−* 0.667) probability of migration.)

| Deme | GPSC | Parameter | Relative Parameter | Directional Migration Probability | ESS |
| --- | --- | --- | --- | --- | --- |
| sa_mal | 10 | 1.542[1.365-1.747] | 1.451[1.285-1.644] | 0.667 | 13594.51 |
| sa_mal | 2 | 1.352[1.184-1.549] | 1.843[1.614-2.111] | 0.667 | 10837.93 |
| sa_mal | 22 | 0.923[0.787-1.084] | 2.215[1.888-2.602] | 0.666 | 12514.85 |
| sa_mal | 26 | 1.033[0.9-1.195] | 1.683[1.467-1.947] | 0.667 | 15763.81 |
| sa_mal | 5 | 1.851[1.655-2.075] | 2.286[2.044-2.562] | 0.667 | 11029.98 |
| sa_mal | 8 | 0.927[0.8-1.082] | 2.492[2.151-2.908] | 0.667 | 21707.4 |
| sa_ken | 10 | 1.211[0.007-1.472] | 1.14[0.007-1.385] | 0.507 | 50.63545 |
| sa_ken | 2 | 0.462[0.004-0.599] | 0.629[0.006-0.816] | 0.411 | 982.8784 |
| sa_ken | 22 | 0.659[0.006-0.829] | 1.581[0.015-1.99] | 0.496 | 145.9672 |
| sa_ken | 26 | 0.507[0.005-0.657] | 0.827[0.008-1.07] | 0.479 | 80.22054 |
| sa_ken | 5 | 0.902[0.016-1.075] | 1.114[0.02-1.328] | 0.577 | 113.1349 |
| sa_ken | 8 | 0.286[0.002-0.372] | 0.77[0.005-1.001] | 0.424 | 247.5898 |
| sa_gam | 10 | 0.547[0.399-0.665] | 0.515[0.375-0.626] | 0.627 | 6574.967 |
| sa_gam | 2 | 1.142[0.993-1.315] | 1.557[1.354-1.792] | 0.667 | 24110.21 |
| sa_gam | 22 | 0.807[0.684-0.951] | 1.938[1.642-2.283] | 0.665 | 23602.96 |
| sa_gam | 26 | 2.953[2.808-2.998] | 4.814[4.578-4.887] | 0.64 | 10354.19 |
| sa_gam | 5 | 1.292[1.146-1.464] | 1.595[1.415-1.808] | 0.667 | 21663.93 |
| sa_gam | 8 | 0.807[0.684-0.953] | 2.17[1.839-2.561] | 0.664 | 18415.62 |
| mal_ken | 10 | 0.066[0.003-0.294] | 0.062[0.002-0.276] | 0 | 12223.04 |
| mal_ken | 2 | 0.064[0.002-0.308] | 0.088[0.003-0.42] | 0 | 10179.53 |
| mal_ken | 22 | 0.068[0.002-0.686] | 0.162[0.005-1.647] | 0.024 | 615.344 |
| mal_ken | 26 | 0.08[0.003-0.75] | 0.131[0.005-1.222] | 0.021 | 1909.27 |
| mal_ken | 5 | 0.063[0.002-0.285] | 0.077[0.003-0.352] | 0 | 11555.99 |
| mal_ken | 8 | 0.069[0.003-0.624] | 0.185[0.007-1.679] | 0.097 | 826.4831 |
| mal_gam | 10 | 1.144[0.139-1.384] | 1.077[0.131-1.302] | 0.638 | 514.1442 |
| mal_gam | 2 | 2.878[2.51-2.995] | 3.921[3.419-4.081] | 0.454 | 6888.511 |
| mal_gam | 22 | 1.53[1.296-1.807] | 3.672[3.111-4.338] | 0.666 | 5317.546 |
| mal_gam | 26 | 2.518[2.184-2.907] | 4.105[3.559-4.738] | 0.667 | 6652.609 |
| mal_gam | 5 | 2.846[2.21-2.994] | 3.515[2.729-3.698] | 0.529 | 1395.229 |
| mal_gam | 8 | 2.224[1.943-2.565] | 5.978[5.223-6.896] | 0.667 | 9834.932 |
| gam_ken | 10 | 0.146[0.006-0.785] | 0.137[0.005-0.739] | 0.047 | 6380.046 |
| gam_ken | 2 | 0.103[0.004-0.506] | 0.14[0.005-0.689] | 0.002 | 9089.028 |
| gam_ken | 22 | 0.101[0.004-0.609] | 0.241[0.009-1.461] | 0.048 | 5777.879 |
| gam_ken | 26 | 0.109[0.004-0.526] | 0.178[0.007-0.857] | 0.011 | 8371.036 |
| gam_ken | 5 | 0.099[0.004-0.426] | 0.123[0.005-0.526] | 0 | 12688.59 |
| gam_ken | 8 | 0.096[0.003-0.559] | 0.259[0.009-1.503] | 0.005 | 5378.5 |
| mal_sa | 10 | 0.127[0.004-1.702] | 0.119[0.004-1.602] | 0.333 | 13594.51 |
| mal_sa | 2 | 0.131[0.004-1.483] | 0.178[0.006-2.02] | 0.333 | 10837.93 |

Table S1 – continued from previous page
| Deme | GPSC | Parameter | Relative Parameter | Directional Migration Probability | ESS |
| --- | --- | --- | --- | --- | --- |
| mal_sa | 22 | 0.127[0.004-1.044] | 0.305[0.009-2.507] | 0.334 | 12514.85 |
| mal_sa | 26 | 0.131[0.004-1.141] | 0.213[0.007-1.861] | 0.333 | 15763.81 |
| mal_sa | 5 | 0.13[0.004-2.032] | 0.161[0.005-2.509] | 0.333 | 11029.98 |
| mal_sa | 8 | 0.121[0.004-1.049] | 0.324[0.01-2.82] | 0.333 | 21707.4 |
| ken_sa | 10 | 0.133[0.004-1.505] | 0.125[0.003-1.416] | 0.493 | 50.63545 |
| ken_sa | 2 | 0.397[0.002-0.616] | 0.541[0.003-0.839] | 0.589 | 982.8784 |
| ken_sa | 22 | 0.115[0.003-0.846] | 0.275[0.006-2.03] | 0.504 | 145.9672 |
| ken_sa | 26 | 0.092[0.002-0.664] | 0.15[0.003-1.082] | 0.521 | 80.22054 |
| ken_sa | 5 | 0.084[0.002-1.082] | 0.104[0.003-1.336] | 0.423 | 113.1349 |
| ken_sa | 8 | 0.13[0.002-0.385] | 0.348[0.005-1.036] | 0.576 | 247.5898 |
| gam_sa | 10 | 0.259[0.004-0.65] | 0.244[0.004-0.611] | 0.373 | 6574.967 |
| gam_sa | 2 | 0.171[0.005-1.273] | 0.233[0.007-1.734] | 0.333 | 24110.21 |
| gam_sa | 22 | 0.203[0.006-0.919] | 0.487[0.014-2.206] | 0.335 | 23602.96 |
| gam_sa | 26 | 2.374[0.053-2.995] | 3.869[0.086-4.882] | 0.36 | 10354.19 |
| gam_sa | 5 | 0.164[0.005-1.411] | 0.202[0.006-1.743] | 0.333 | 21663.93 |
| gam_sa | 8 | 0.185[0.005-0.905] | 0.498[0.013-2.434] | 0.336 | 18415.62 |
| ken_mal | 10 | 1.588[0.008-1.865] | 1.494[0.007-1.755] | 1 | 12223.04 |
| ken_mal | 2 | 1.054[0.007-1.28] | 1.436[0.01-1.744] | 1 | 10179.53 |
| ken_mal | 22 | 0.537[0.012-0.713] | 1.289[0.028-1.712] | 0.976 | 615.344 |
| ken_mal | 26 | 0.645[0.009-0.835] | 1.052[0.014-1.362] | 0.979 | 1909.27 |
| ken_mal | 5 | 1.534[0.008-1.816] | 1.894[0.01-2.242] | 1 | 11555.99 |
| ken_mal | 8 | 0.361[0.005-0.56] | 0.969[0.013-1.505] | 0.903 | 826.4831 |
| gam_mal | 10 | 0.241[0.007-1.408] | 0.227[0.007-1.325] | 0.362 | 514.1442 |
| gam_mal | 2 | 2.797[1.892-2.993] | 3.812[2.577-4.077] | 0.546 | 6888.511 |
| gam_mal | 22 | 0.191[0.006-1.714] | 0.457[0.013-4.114] | 0.334 | 5317.546 |
| gam_mal | 26 | 0.232[0.007-2.826] | 0.379[0.011-4.606] | 0.333 | 6652.609 |
| gam_mal | 5 | 2.512[0.198-2.988] | 3.102[0.244-3.69] | 0.471 | 1395.229 |
| gam_mal | 8 | 0.209[0.007-2.484] | 0.562[0.017-6.679] | 0.333 | 9834.932 |
| ken_gam | 10 | 0.62[0.015-0.822] | 0.584[0.014-0.774] | 0.953 | 6380.046 |
| ken_gam | 2 | 0.966[0.012-1.18] | 1.316[0.017-1.608] | 0.998 | 9089.028 |
| ken_gam | 22 | 0.549[0.011-0.726] | 1.319[0.027-1.742] | 0.952 | 5777.879 |
| ken_gam | 26 | 0.712[0.018-0.897] | 1.161[0.03-1.462] | 0.989 | 8371.036 |
| ken_gam | 5 | 1.335[0.016-1.593] | 1.648[0.019-1.968] | 1 | 12688.59 |
| ken_gam | 8 | 0.753[0.012-0.932] | 2.023[0.031-2.506] | 0.995 | 5378.5 |

**Table S2:** Migration parameter estimates symmetrically across four demes for six GPSCs. Values within the square brackets denote the 95% confidence intervals. The ‘Parameter’ is the raw migration parameter estimate while the ‘Relative Parameter’ is relative to all other deme pairs *within* each GPSC.

| Deme | GPSC | Parameter | Relative Parameter |
| --- | --- | --- | --- |
| sa_mal | 10 | 1.761[0.241-4.655] | 1.031[0.141-2.725] |
| sa_ken | 10 | 0.841[0.064-1.916] | 0.492[0.037-1.121] |
| sa_gam | 10 | 3.156[0.57-4.862] | 1.847[0.333-2.846] |
| mal_ken | 10 | 1.606[0.106-4.751] | 0.94[0.062-2.781] |
| mal_gam | 10 | 1.336[0.253-2.863] | 0.782[0.148-1.676] |
| gam_ken | 10 | 1.119[0.186-2.847] | 0.655[0.109-1.666] |
| sa_mal | 2 | 1.465[0.108-3.99] | 0.833[0.061-2.268] |
| sa_ken | 2 | 2.135[0.267-3.816] | 1.213[0.152-2.169] |
| sa_gam | 2 | 0.886[0.076-2.675] | 0.504[0.043-1.52] |
| mal_ken | 2 | 0.138[0.005-0.961] | 0.078[0.003-0.546] |
| mal_gam | 2 | 4.242[3.047-4.937] | 2.411[1.732-2.806] |
| gam_ken | 2 | 1.476[0.462-2.383] | 0.839[0.262-1.354] |
| sa_mal | 22 | 1.755[0.121-4.77] | 1.077[0.074-2.927] |
| sa_ken | 22 | 2.111[0.204-4.721] | 1.295[0.125-2.897] |
| sa_gam | 22 | 0.726[0.023-2.728] | 0.445[0.014-1.674] |
| mal_ken | 22 | 1.258[0.063-4.492] | 0.772[0.039-2.757] |
| mal_gam | 22 | 1.777[0.109-4.608] | 1.091[0.067-2.828] |
| gam_ken | 22 | 0.863[0.03-3.116] | 0.53[0.019-1.912] |
| sa_mal | 26 | 0.805[0.042-3.071] | 0.473[0.025-1.802] |
| sa_ken | 26 | 4.07[2.368-4.948] | 2.388[1.39-2.903] |
| sa_gam | 26 | 0.779[0.032-2.133] | 0.457[0.019-1.251] |
| mal_ken | 26 | 0.464[0.015-2.029] | 0.272[0.009-1.191] |
| mal_gam | 26 | 2.777[1.818-4.105] | 1.63[1.067-2.409] |
| gam_ken | 26 | 1.027[0.082-2.095] | 0.602[0.048-1.229] |
| sa_mal | 5 | 1.42[0.178-4.208] | 0.684[0.086-2.028] |
| sa_ken | 5 | 1.935[0.679-4.668] | 0.933[0.327-2.25] |
| sa_gam | 5 | 1.45[0.088-4.149] | 0.699[0.042-2] |
| mal_ken | 5 | 0.437[0.018-1.476] | 0.211[0.009-0.712] |
| mal_gam | 5 | 4.544[3.209-4.986] | 2.19[1.547-2.404] |
| gam_ken | 5 | 1.925[0.698-4.79] | 0.928[0.337-2.309] |
| sa_mal | 8 | 1.446[0.086-4.485] | 0.872[0.052-2.705] |
| sa_ken | 8 | 2.05[0.138-4.818] | 1.236[0.083-2.905] |
| sa_gam | 8 | 0.74[0.037-2.669] | 0.446[0.022-1.609] |
| mal_ken | 8 | 0.93[0.038-4.051] | 0.561[0.023-2.443] |
| mal_gam | 8 | 2.585[0.481-4.835] | 1.559[0.29-2.916] |
| gam_ken | 8 | 1.042[0.051-4.051] | 0.628[0.031-2.443] |

